# Mapping Neurochemical Signatures onto Brain Structure for Neurotransmitter-Informed Discrimination of Schizophrenia Patients from Healthy Controls

**DOI:** 10.64898/2025.12.04.25341533

**Authors:** Lisa Hahn, Florian J. Raabe, Daniel Keeser, Clara Vetter, John Fanning, Alkomiet Hasan, Irina Papzova, Ariane Wiegand, Peter Falkai, Eva Meisenzahl, Nikolaos Koutsouleris

## Abstract

**Background:** Schizophrenia (SCZ) is associated with widespread gray matter volume (GMV) reductions, yet the underlying mechanisms driving these alterations remain unclear. This study investigates whether SCZ-related GMV changes co-localize with normative neurotransmitter (NT) density maps derived from healthy individuals. We further examine the diagnostic utility and transdiagnostic relevance of these NT-informed structural patterns within a machine learning (ML) framework.

**Methods:** Structural imaging from 445 SCZ patients and 414 matched healthy controls (HC) were used to train two ML models: (1) an NT-informed model based on spatial cortical and subcortical correlations between GMV and 25 normative NT maps, and (2) an ‘NT-unaware’ control model using 50 principal components of GMV. Both models were tested for transdiagnostic generalizability in major depression (MDD), bipolar disorder (BD), borderline personality disorder (BPD), and attention-deficit/hyperactivity disorder (ADHD) to determine whether NT-informed and NT-uninformed morphometric patterns reflect psychosis-specific mechanisms. Associations with accelerated brain aging, clinical symptoms, and psychotropic medication exposure were evaluated. Individual feature importance profiles from the NT model were used for clustering to identify potential neurochemical subtypes of SCZ.

**Results:** The NT model had a higher sensitivity, but a lower specificity compared to the control model in distinguishing SCZ from HC (sensitivity_NT_ = 61.8% vs. sensitivity_PCA_ = 58.4%; specificity_NT_ = 62.3% vs. specificity_PCA_ = 81.9%). The NT model’s SCZ-predictive features localized primarily to subcortical regions and mapped onto serotonergic (5-HT4), endocannabinoid (CB1), and cholinergic (VAChT) systems, as well as synaptic (SV2A) and gene expression regulation (HDAC) markers. In contrast, the control model relied on medial temporal and subcortical structures, and its decision scores were associated with accelerated brain aging (R^2^ = 0.11). The NT model showed modest transfer to BPD (BAC = 61.8%, p = 0.003) and MDD (BAC = 56.6%, p = 0.036), while the control model performed better for MDD (BAC = 61.2%, p < 0.001) and BD (BAC = 61.0%, p = 0.002). Finally, clustering of individual NT feature contributions identified two transdiagnostic NT subtypes: a subcortical dopaminergic/COX-1/mGluR5-driven subtype and a subtype with diffuse cortical and subcortical serotonergic-GABAergic-glutamatergic-cholinergic involvement.

**Conclusions:** Spatial correlations with normative NT distributions provide biologically interpretable insights into SCZ-related GMV alterations, particularly within subcortical systems. The NT-informed model identifies mechanistically distinct subtypes potentially relevant for future NT-based stratification of SCZ. These findings support the future integration of molecular and structural imaging to uncover system-specific vulnerabilities in psychosis and guide more personalized approaches to diagnosis and treatment.

## Introduction

Schizophrenia (SCZ) is a complex psychiatric condition with substantial heterogeneity in both clinical presentation and underlying neurobiology ^1–3^. This variability presents a major challenge for treatment development and underscores the importance of understanding the mechanisms driving brain alterations in SCZ. Historically, the dopamine hypothesis of regional specificity has been central to conceptualizing SCZ pathophysiology, proposing that mesolimbic dopaminergic hyperactivity underlies positive symptoms, while prefrontal cortical hypodopaminergia contributes to negative and cognitive symptoms ^4–9^. This framework has guided antipsychotic drug development, particularly those targeting dopamine D2 receptors (D2). However, its limitations are increasingly recognized, as it fails to explain the large variety of clinical and biological features observed in SCZ ^10,11^. Accumulating evidence implies dysfunction in additional neurotransmitter (NT) systems, for example glutamatergic hypofunction and particularly N-methyl-D-aspartate (NMDA) receptor dysfunction has been linked to negative symptoms and cognitive deficits ^12,13^. Furthermore, gamma-aminobutyric acid (GABA)-ergic abnormalities are thought to be associated with disrupted cortical synchrony ^14,15^. Serotonergic and cholinergic systems have also been implicated in SCZ pathophysiology, as supported by pharmacological and meta-analytic evidence ^16,17^.

From a neuroanatomical perspective, SCZ is underpinned by regionally specific gray matter volume (GMV) reductions, particularly in the anterior cingulate cortex, superior temporal gyrus, medial prefrontal cortex, insula, hippocampus, thalamus, and amygdala ^18–21^. Recent meta-analytic evidence further shows that gray matter (GM) patterns allow above-chance diagnostic classification of SCZ ^22^. Similar regions have also been implicated in elevated Brain Age Gap Estimation (BrainAGE) defined as the difference between predicted and chronological age, a phenomenon observed in patients with SCZ and other psychiatric conditions, which is associated with poorer clinical outcomes ^23,24^. A recent transdiagnostic systematic review synthesized magnetic resonance imaging (MRI)-derived BrainAGE findings across SCZ and other psychotic disorders, bipolar disorder (BD), mood disorders such as major depression (MDD), and stress-related disorders, highlighting shared BrainAGE topographies ^25^. Moreover, several major psychiatric disorders share overlapping neurobiological features, including structural abnormalities and NT system dysfunction ^26–28^, pointing to partial transdiagnostic convergence of structural and NT system alterations.

While these structural changes are well-documented, their molecular and cellular underpinnings remain unclear. Recent studies provide good evidence that GMV volume reduction is driven by a loss of synaptic structures. However, it is unclear whether this synaptic loss particularly effects specific NT systems on the molecular level. Recent advances in NT-informed neuroimaging offer new ways to explore this link by combining GMV data with normative (i.e., derived from healthy volunteer populations) positron emission tomography (PET)- and single-photon emission computed tomography (SPECT)-derived maps of receptor and transporter density. Tools such as JuSpace ^29^ and Neuromaps ^30^ enable spatial correlation analyses across major NT systems, and recent work has shown that GMV alterations in SCZ co-localize with high serotonin transporter (5-HTT) densities, suggesting spatially organized, system-specific vulnerability ^31^. In contrast, a different study found positive associations with D2, the dopamine transporter (DAT), and norepinephrine transporter (NET), and negative associations with serotine 4 receptor (5-HT4) and cannabinoid receptor type 1 (CB1) ^32^. These findings suggest biological heterogeneity of the NT systems underlying macroscopic neuroanatomical signatures of the disorder, which may explain the variability in its clinical presentation.

Building on prior findings, we hypothesize that SCZ-related GM pathology spatially overlaps with normative density maps of key NT systems. Our primary aim is to test whether these alterations preferentially affect regions enriched for dopaminergic, serotonergic, glutamatergic, and GABAergic receptors and transporters, indicating system-specific vulnerability of synaptic disturbances in SCZ. To test this, we integrate normative NT density maps included in Neuromaps ^30^ with GMV as derived from structural MRI (sMRI) using a spatial co-localization approach together with a machine learning (ML) classification framework separating SCZ patients from healthy controls (HC). In addition to an NT-informed classifier, we implemented a principal component analysis (PCA)-based, NT-agnostic control model derived solely from GMV, allowing us to compare their classification performance and predicted phenotypic profiles. Here, ML is used to capture multivariate patterns and assess predictive generalizability. To evaluate transdiagnostic generalizability, we extend our analyses to MDD, BD, borderline personality disorder (BPD), and attention-deficit/hyperactivity disorder (ADHD), thereby assessing whether NT-informed and NT-uninformed morphometric patterns capture psychosis-specific mechanisms or broader, shared neurobiological vulnerability across psychiatric disorders. To further examine these patterns, we explore their association with BrainAGE, allowing us to test whether NT-related structural vulnerability not only supports diagnostic classification but also reflects individual variability in age-related brain trajectories. Finally, to assess whether distinct NT co-localization patterns account for variability across individuals and diagnostic categories, we explore mechanistic subtypes by clustering based on NT-specific feature importance.

## Methods

### Participants and sMRI processing

We included 445 patients with SCZ (mean age = 34.0 ± 11.3 years; 103 females) and 847 HC (mean age = 33.3 ± 11.1 years; 368 females) in the current study. To ensure comparability across sites, 414 HCs (mean age = 34.3 ± 12.1 years; 185 females) were matched to SCZ patients by age and sex across sites. The remaining 433 HCs together with remaining patient groups including MDD (N = 104; mean age = 42.3 ± 12.0 years; 52 females), BD (N = 101; mean age = 39.2 ± 11.0 years; 48 females), ADHD (N = 41; mean age = 32.3 ± 10.4 years; 20 females), and BPD (N = 59; mean age = 25.9 ± 7.0 years; 59 females) were selected for transdiagnostic comparisons. All participants were pooled from five independent cohorts including the MIND Clinical Imaging Consortium (MCIC) ^33^, the Center for Biomedical Research Excellence (COBRE) ^34^, the UCLA Consortium for Neuropsychiatric Phenomics LA5c Study (UCLA) ^35^, the Multimodal Imaging in Chronic Schizophrenia Study (MIMICSS; part of the PsyCourse study) ^36–38^, and the Munich Imaging Database (MUC) ^23,39^. The detailed distribution of diagnostic groups including demographics across sites and cohorts is reported in Supplementary Table S1 and Supplementary Figure S1. Written informed consent was collected from each participant. The study was approved by the local ethics committees of the respective universities for each cohort and in accordance with the Declaration of Helsinki.

Structural T1-weighted magnetization-prepared rapid gradient-echo (MPRAGE) MRI was acquired on 1.5T and 3T scanners (see Supplementary Table S2 for site-specific imaging parameters). To calculate voxel-wise GMV, structural images were segmented, spatially normalized to MNI space, and modulated using the standalone CAT12.8 version 1933 (https://neuro-jena.github.io/enigma-cat12/). In the case of the availability of multiple visits per participant, only the first visit was considered for analysis (see Supplementary Methods for information on MRI data acquisition, preprocessing and covariate correction). This resulted in the exclusion of 13 subjects from the COBRE cohort since they were part of the preceding MCIC cohort.

### Support vector classification and transdiagnostic application

We constructed two unimodal classifiers, an NT-informed model and a PCA-based, NT-agnostic, purely structural-imaging-based control model, to differentiate SCZ from HC and compare both classification accuracy and predicted phenotypic profiles. Classification was performed on covariate-corrected (i.e., for age, sex and site) GMV maps within a repeated nested leave-one-site-out cross-validation (CV) framework. For the NT-informed model, features reflected cortical and subcortical partial correlations between GMV and 25 normative NT density maps included in Neuromaps (30; see Supplementary Table S4 for details), whereas the control model applied PCA restricted to 50 principal components to match feature dimensionality (see Supplementary Methods for details about the computation of the NT correlations). Model training, feature selection, and evaluation were performed using linear support vector machines with balanced accuracy (BAC) as the primary optimization criterion. Details on preprocessing, nested CV structure, and statistical comparisons between models are provided in the Supplementary Methods (see Support vector classification).

Next, we assessed the similarity between the two models’ decision scores using Spearman correlation and evaluated model performance significance through permutation testing. Furthermore, we used McNemar’s test to determine whether the two classifiers differed significantly in their error patterns. Feature relevance and stability were evaluated using CV ratio (CVR) mapping at a stability threshold of ≥ |2|, filtered by significant, false discovery rate (FDR) corrected sign-based consistency values (pFDR < 0.05). CVR was computed as the sum of the selected median weights from the inner CV1 folds across all CV2 folds, divided by the standard error of these weights for each feature, whereas sign-based consistency reflects how reliably a feature’s weight maintains the same direction (i.e., positive or negative) across CV folds. Both visualization methods are described in detail in the supplementary material of Koutsouleris et al. (2021) ^40^. To infer the directionality of the GMV features of the PCA model, a threshold-free cluster enhancement (TFCE) analysis with 5,000 permutations was performed using the TFCE toolbox (https://github.com/ChristianGaser/tfce) on the covariate-corrected images, applying FDR correction at α = 0.05 and restricting the analysis to the CVR map as a mask to identify GMV increases (SCZ > HC) and decreases (HC > SCZ). Moreover, to infer the directionality of the correlation coefficients, z-score maps of each patient relative to HC were computed within each CV2 fold and then correlated with the normative NT distributions (see Directional Inference via Cross-Validated Neurotransmitter Correlation in the Supplementary Methods). Additional analyses were conducted to assess residual sociodemographic biases on decision scores. Detailed descriptions of these procedures are provided in the Supplementary Methods.

Finally, the transdiagnostic generalizability of both classifiers was evaluated by applying the models to the remaining patient groups present in the five cohorts including patients with MDD, BD, ADHD, and BPD (see Supplementary Table S1). The significance of these model applications was assessed using permutation testing with label shuffling.

### Post hoc association with BrainAGE, clinical symptoms and psychotropic medication exposure

To assess the influence of BrainAGE on both models’ performance, BrainAGE was computed using a ν-support vector regression model trained on all HC (N = 847) within the same nested CV design. The preprocessing and model training procedures, along with age-bias correction methods, are detailed in the Supplementary Methods. The model was applied to all patient groups to derive BrainAGE scores as the difference between predicted and chronological age.

A two-way analysis of covariance (ANCOVA) was conducted to examine the effects of Diagnosis (six groups: SCZ, MDD, ADHD, BD, BPD) and BrainAGE (continuous covariate) including their interaction on the decision scores of both models. Analyses were conducted in R (version 4.5.0) using the lm() function and post-hoc simple slopes or marginal means were estimated using the emmeans package (emtrends() and emmeans() functions). To assess the strength of the association between BrainAGE and decision scores within each diagnostic group, we computed the explained variance (η²) from separate simple linear regressions of BrainAGE on the models’ decision scores.

Next, to examine if the decision scores were associated with a specific, potentially transdiagnostic phenotype, separate linear regression models (LMs) were computed between decision scores and Positive and Negative Syndrome Scale (PANSS) ^41^ factors (positive, negative, disorganized/concrete, excited, depressed) according to Wallwork and colleagues ^42^, global ratings of the Scale for the Assessment of Positive Symptoms (SAPS) ^43^ and Scale for the Assessment of Negative Symptoms (SANS) ^44^. For illness duration and depressive scales including the Calgary Depression Scale for Schizophrenia (CDSS) ^45^ and the Hamilton Depression Rating Scale (HDRS-17) ^46^, Spearman correlation coefficients were computed between scale total scores and decision scores of both models. Finally, additional LMs were computed at the item level (excluding factors, global ratings, and total scores) for all available symptom scales, and tolerance (computed as 1/variance inflation factor) was inspected to assess potential multicollinearity among predictors. Given the exploratory nature of this analysis, only medium effects as measured by the effect size η² (i.e., η² ≥ 0.06) were considered.

Finally, to examine whether psychotropic medication exposure was associated with model predictions, we compared decision scores between drug-naïve and exposed patients for antipsychotics, antidepressants, benzodiazepines, and mood stabilizers computing separate LMs per cohort (see Supplementary Table S13). Additionally, the effect size η² was computed to evaluate the explained variance for each medication category. Details on the availability of medication information for each cohort are reported in the Supplementary Methods.

### Clustering individual feature importance of the NT model and comparing BrainAGE and clinical symptoms between clusters

We first quantified feature contributions at the subject level (i.e., on case-by-case level) by applying median-flip perturbation (i.e., shifting percentiles by ±50 across the median) to each subject’s NT-correlation features, yielding one feature importance profile per individual. To identify mechanistic subtypes among individuals classified as patients, we clustered individual feature importance profiles using unsupervised ML. Details on the median flip procedure, considered clustering algorithms, and validation metrics are reported in the Supplementary Methods and Supplementary Table S14. The optimal and most stable solution was achieved with k-means clustering (k = 2), revealing two robust clusters comprising 237 and 497 individuals, respectively. To examine potential differences regarding cluster assignments, a chi-square test of independence was performed between cluster assignment and label (i.e., original patient or HC). To visualize differences in feature importance of the NT model between clusters, we statistically compared the distributions using Wilcoxon rank-sum tests, adjusting for multiple comparisons across NTs using FDR-correction, and computed the effect size r as standardized z-values (i.e., 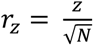).

To assess differences in cluster compactness, we calculated each participant’s Euclidean distance to their respective cluster centroid and statistically compared this distance between clusters using a Wilcoxon rank-sum test. Additionally, we computed separate LMs per cluster and scale to assess whether there was a relationship between distance to centroid and PANSS factors, and global ratings of SAPS and SANS. For the HDRS-17 total score and illness duration, Spearman correlation coefficients were computed instead.

Finally, to evaluate differences in BrainAGE, demographics including age and sex, illness duration, PANSS factors, global ratings of SAPS and SANS, and total scores measuring depressive symptoms between clusters, we conducted Wilcoxon rank-sum tests for continuous variables and chi-square tests of independence for categorical variables. FDR correction was applied to adjust for multiple comparisons within each scale.

## Results

### Support-vector classification and transdiagnostic application

The NT model utilizing cortical and subcortical spatial correlations between GMV and 25 normative NT density maps from independent healthy volunteer populations performed with a BAC of 62.1% (sensitivity = 61.8%, specificity = 62.3%, *p* = 0.027; see Figure 1A). In contrast, the purely GMV-based PCA model utilizing 50 GMV-derived principal components had a lower sensitivity of 58.4%, but higher specificity of 81.9% (BAC = 70.2%; *p* < 0.001; see Figure 1B). Additional performance measures are reported in Table 1.

**Figure 1.**
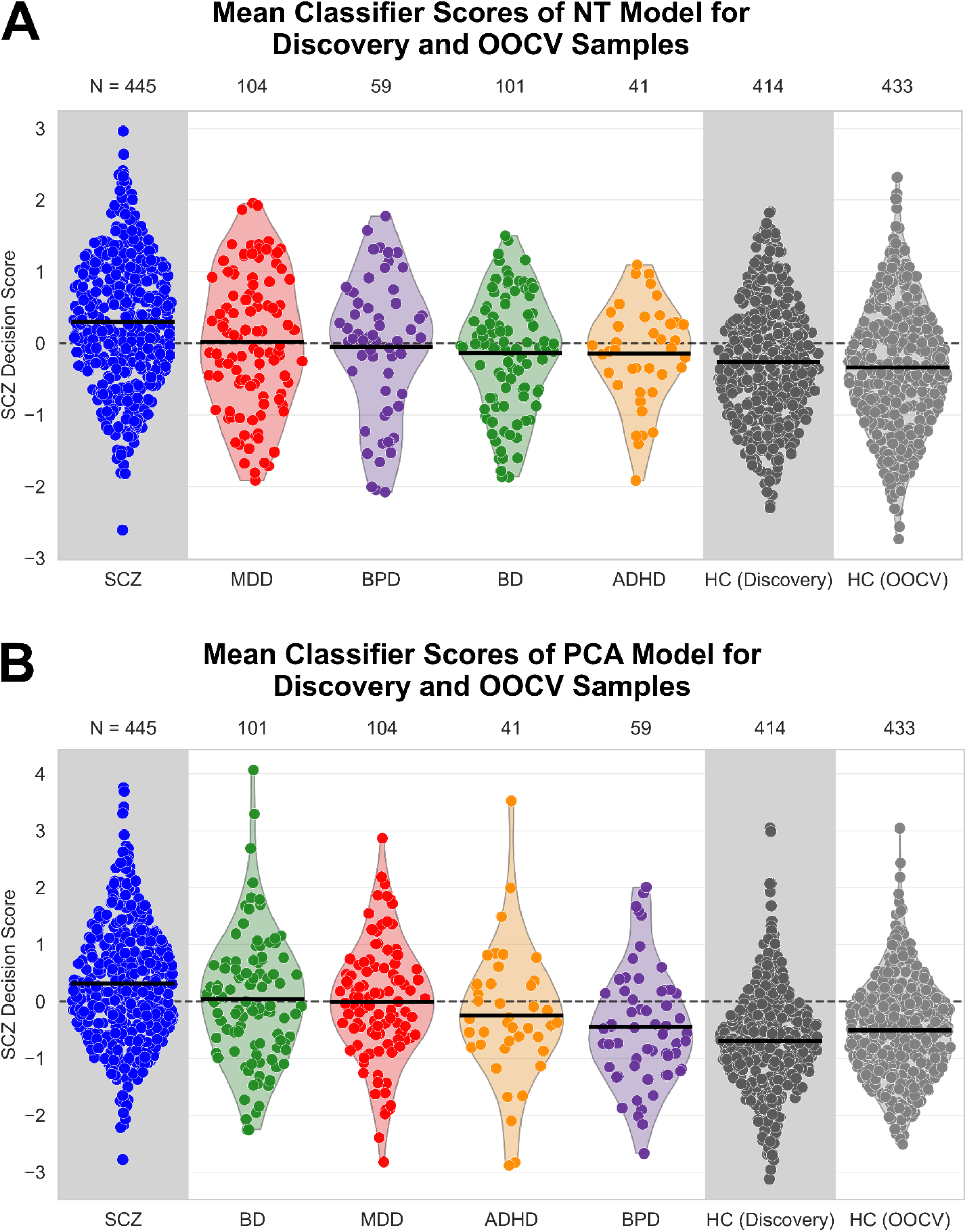
Mean decision scores of NT and PCA models for discovery and OOCV samples. Mean decision scores of **(A)** NT model and **(B)** PCA model for SCZ vs. HC classification and the transdiagnostic application to MDD, BD, BPD, and ADHD.

**Table 1.** Performance of NT and PCA models for the classification of SCZ and HC, and the application to BD, MDD, ADHD, and BPD.

|  | **Classification** | **BAC** | **Sens** | **Spec** | **AUC (95-CI)** | ***p*** |
| --- | --- | --- | --- | --- | --- | --- |
| NT | HC-SCZ | 62.1% | 61.8% | 62.3% | 0.67<br>(0.64-0.71) | 0.027 |
|  | HC-MDD | 56.6% | 49.0% | 64.2% | 0.61<br>(0.54-0.67) | 0.036 |
|  | HC-BD | 53.9% | 43.6% | 64.2% | 0.58<br>(0.51-0.64) | 0.081 |
|  | HC-ADHD | 55.3% | 46.3% | 64.2% | 0.58<br>(0.49-0.68) | 0.071 |
|  | HC-BPD | 61.8% | 59.3% | 64.2% | 0.60<br>(0.52-0.68) | 0.003 |
| PCA | HC-SCZ | 70.2% | 58.4% | 81.9% | 0.77<br>(0.74-0.80) | < 0.001 |
|  | HC-MDD | 61.2% | 49.0% | 73.4% | 0.66<br>(0.60-0.72) | < 0.0001 |
|  | HC-BD | 61.0% | 48.5% | 73.4% | 0.65<br>(0.59-0.71) | 0.002 |
|  | HC-ADHD | 55.0% | 36.6% | 73.4% | 0.58<br>(0.49-0.68) | 0.285 |
|  | HC-BPD | 52.0% | 30.5% | 73.4% | 0.51<br>(0.43-0.59) | 0.384 |
| Stacked | HC-SCZ | 69.5% | 57.8% | 81.2% | 0.77<br>(0.74-0.80) | --- |

ADHD – Attention Deficit Hyperactivity Disorder, AUC – Area Under the Curve, BAC – Balanced Accuracy, BD – Bipolar Disorder, BPD – Borderline Personality Disorder, MDD – Major Depression, SCZ – Schizophrenia, Sens – Sensitivity, Spec – Specificity

The resulting CVR maps revealed stable predictive features distinguishing between SCZ and HC. For the NT model, SCZ classification was associated exclusively with subcortical regions, demonstrating positive correlations (i.e., SCZ patients showing increased GMV relative to HC) with 5-HT4, CB1, histone deacetylase (HDAC), synaptic vesicle glycoprotein 2A (SV2A), and the vesicular acetylcholine transporter (VAChT; see Figure 2A and Supplementary Figures S4A and S4B). In contrast, HC classification was associated with a positive cortical correlation for cyclooxygenase-1 (COX-1) and a negative cortical correlation (i.e., SCZ patients showing decreased GMV relative to HC) for CB1 and positive subcortical correlations for D2 and DAT, and negative subcortical correlations for serotonin 1A receptor (5-HT1A) and GABA receptor α (GABAA). For the PCA model, SCZ classification was mainly associated with subcortical structures including bilateral basal ganglia structures, parts of the thalamus, and medial temporal areas, whereas HC classification was predominantly localized in frontal, temporal, parietal, and occipital cortices (see Figure 2B). Consistent with this pattern, the TFCE permutation analysis revealed lower GMV in SCZ across widespread frontal, parietal, temporal, and occipital cortical regions, as well as the cerebellum and brainstem, while higher GMV in schizophrenia was observed in the basal ganglia (putamen, pallidum), thalamus, and adjacent medial temporal/limbic regions (see Supplementary Figure S2).

**Figure 2.**
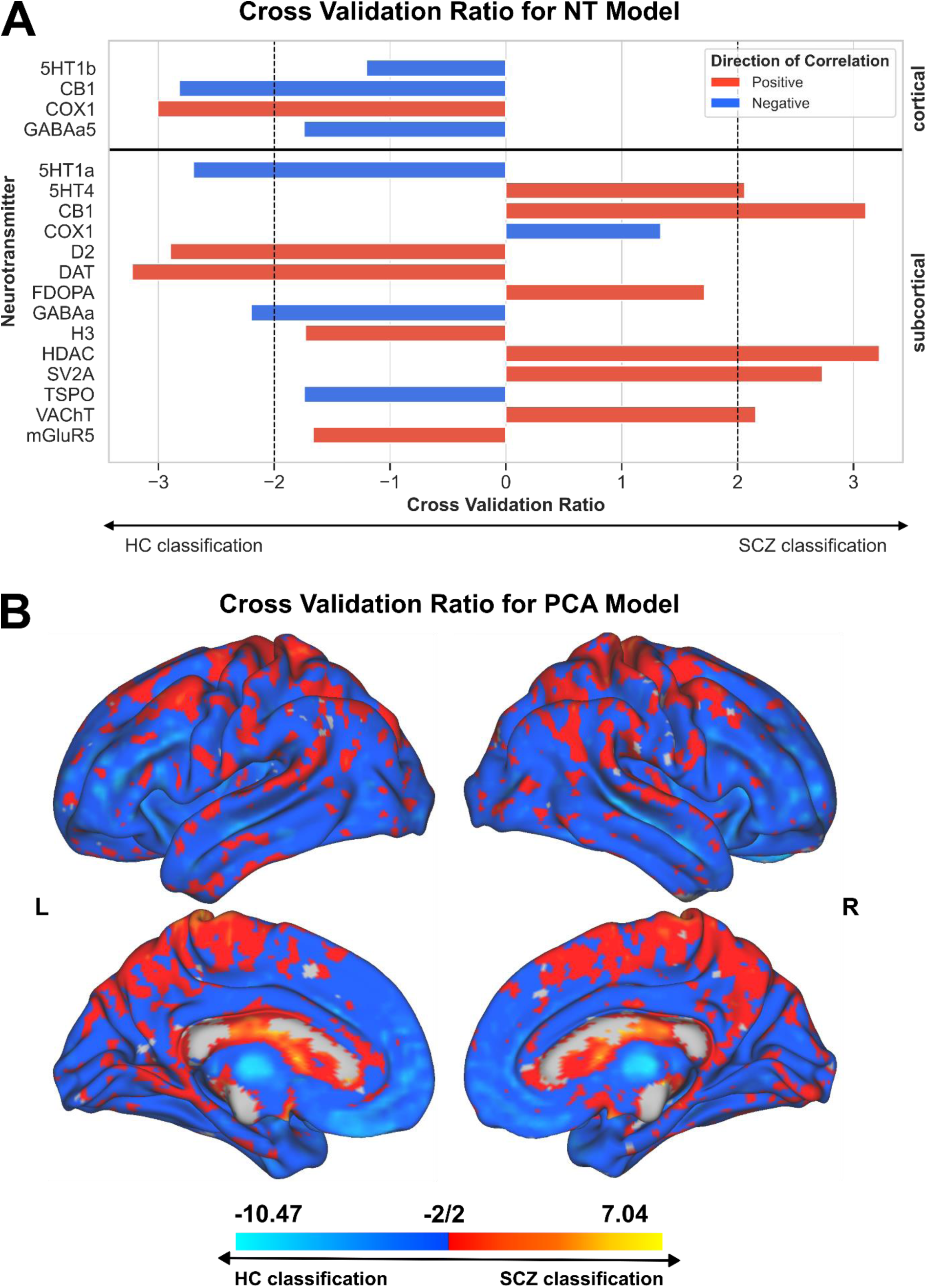
Cross validation ratios for NT and PCA models. Cross-validation ratios were filtered for FDR-adjusted sign-based consistency values for the **(A)** NT model and **(B)** PCA model.

Evaluating the alignment between models, the Spearman correlation between the decision scores showed a medium association strength between the models (*r* = 0.60, *p* < 0.0001, *R²* = 0.36), showing that 36% of the variance in the decision scores can be explained by the relationship between the models (see Supplementary Results for statistical comparison of model performance and stacked model). Additionally, McNemar’s test showed a significant asymmetry in misclassifications between the models (χ²(1) = 36.66, p < .001), indicating that their predictions differed substantially. The decision scores of all models (NT, PCA, and stacked) were not significantly influenced by site, age, or sex (all *p* > 0.05), indicating that the applied correction methods effectively controlled for these confounds (see Supplementary Table S5 for statistics and corresponding p-values).

The NT model showed a significant transdiagnostic application performance in BPD (BAC = 61.8%, sensitivity = 59.3%, specificity = 64.2%, *p* = 0.003) and MDD (BAC = 56.6%, sensitivity = 49.0%, specificity = 64.2%, *p* = 0.036), but not in ADHD, or BD (BD: BAC = 53.9%; ADHD: BAC = 55.3%; all *p* > 0.05). In contrast, the PCA model performed best in MDD (BAC = 61.2%, sensitivity = 49.0%, specificity = 73.4%, *p* < 0.001) and BD (BAC = 61.0%, sensitivity = 48.5%, specificity = 73.4%, *p* = 0.002), with non-significant results for ADHD, and BPD (BPD: BAC = 52.0%; ADHD: BAC = 55.0%; all *p* > 0.05). Additional performance metrics are reported in Table 1.

### Post hoc associations of model decisions with BrainAGE, clinical symptoms and psychotropic medication exposure

#### Association with BrainAGE

For the NT model, a two-way ANCOVA revealed that there was no main effect of BrainAGE (*F*(1, 740) = 0.08, *p* = .784), a significant main effect of Diagnosis, *F*(4, 740) = 7.97, *p* < .001) and no significant BrainAGE × Diagnosis interaction (*F*(4, 740) = 1.48, *p* = .208) on the model’s decision scores. Post-hoc FDR-adjusted pairwise comparisons of estimated marginal means indicated that SCZ patients had higher decision scores (i.e., more SCZ-like) than those with ADHD (*t*(740) = 3.00, *p_FDR_* = .026) and BD (*t*(740) = 4.33, *p_FDR_* < .001). No other pairwise differences reached statistical significance after FDR correction (all *p_FDR_* > .05; see Supplementary Table S6).

In contrast, for the PCA model, there were significant main effects of BrainAGE (*F*(1, 740) = 99.80, *p* < .001) and Diagnosis (*F*(4, 740) = 13.79, *p* < .001), as well as a significant BrainAGE × Diagnosis interaction (*F*(4, 740) = 2.81, *p* = .024) on the model’s decision scores. Post-hoc estimated simple slopes (i.e., BrainAGE trends) revealed a positive association of BrainAGE on the model’s decision scores across all groups (see Supplementary Table S6). The strongest association was observed in the ADHD group (*b* = 0.15), followed by BPD (*b* = 0.10), BD (*b* = 0.07), SCZ (*b* = 0.06), and MDD (*b* = 0.05). While FDR-adjusted pairwise comparisons revealed that these slopes did not differ significantly between diagnostic groups (all adjusted *p_FDR_* > .05), exploratory within-group correlations between BrainAGE and decision scores suggested varying degrees of association, with BrainAGE explaining between 5% and 34% of the variance across diagnoses (see Supplementary Table S7). Together, these findings indicate that BrainAGE was unrelated to decision scores in the NT model, whereas the PCA model showed a consistent positive association between higher BrainAGE and more SCZ-like decisions across diagnostic groups.

#### Association with clinical symptoms

Next, we assessed associations of the classifiers’ decision scores with illness duration, positive, negative, and depressive symptoms. Illness duration was not significantly related to decision scores for either model (all p > 0.05, all η² ≤ 0.01). However, there was a significant positive association between the PCA model’s decision scores and the SAPS global rating of hallucinations (*b* = .17, *SE* = .05, *t*(200) = 3.20, *p* = .002, η² = .04), such that greater hallucination severity was associated with more SCZ-like model decisions. Additionally, higher scores on the Wallwork PANSS positive factor were linked to more SCZ-like PCA model decision scores (*b* = .12, *SE* = .06, *t*(325) = 2.13, *p* = .034, η² = .02). Full details are provided in Tables 2 and 3.

**Table 2.** Linear model coefficients (Estimates, SEs, *t*-values, *p*-values, and partial η²) for associations between clinical symptom domains (SAPS, SANS, PANSS) and NT and PCA model decision scores.

|  |  | NT model |  |  |  |  | PCA model |  |  |  |  |
| --- | --- | --- | --- | --- | --- | --- | --- | --- | --- | --- | --- |
| | | Estimate | SE | T | *p* | $\eta^2$ | Estimate | SE | T | *p* | $\eta^2$ |
| SAPS<br>(global) | Intercept | 0.11 | 0.11 | 1.03 | 0.302 | --- | 0.03 | 0.13 | 0.25 | 0.806 | --- |
|  | Hallucinations | 0.07 | 0.04 | 1.59 | 0.114 | 0.01 | 0.17 | 0.05 | 3.20 | 0.002* | 0.04 |
|  | Delusions | 0.02 | 0.04 | 0.45 | 0.656 | 0.00 | -0.05 | 0.05 | -0.96 | 0.338 | 0.00 |
|  | Bizzare Behavior | -0.11 | 0.06 | -1.96 | 0.051 | 0.01 | -0.09 | 0.07 | -1.27 | 0.205 | 0.00 |
|  | Positive Formal Thought Disorder | 0.06 | 0.05 | 1.31 | 0.193 | 0.01 | 0.11 | 0.06 | 1.89 | 0.061 | 0.02 |
| | Model statistics | $F(4, 200) = 1.88, p = 0.116, R^2 = 0.04$ | | | | | $F(4, 200) = 3.38, p = 0.011^*, R^2 = 0.06$ | | | | |
| SANS<br>(global) | Intercept | 0.03 | 0.12 | 0.28 | 0.778 | --- | -0.06 | 0.15 | -0.43 | 0.671 | --- |
|  | Affective Flattening | 0.06 | 0.04 | 1.31 | 0.192 | 0.01 | 0.01 | 0.06 | 0.10 | 0.923 | 0.01 |
|  | Alogia | -0.03 | 0.05 | -0.72 | 0.472 | 0.00 | -0.01 | 0.06 | -0.12 | 0.905 | 0.00 |
|  | Avolition Apathy | 0.04 | 0.05 | 0.91 | 0.361 | 0.00 | 0.09 | 0.06 | 1.56 | 0.120 | 0.02 |
|  | Anhedonia Asociality | -0.02 | 0.05 | -0.52 | 0.600 | 0.00 | 0.07 | 0.06 | 1.26 | 0.210 | 0.01 |
|  | Attention | 0.06 | 0.04 | 1.44 | 0.150 | 0.01 | 0.02 | 0.05 | 0.41 | 0.682 | 0.00 |
| | Model statistics | $F(5, 266) = , p = 0.358, R^2 = 0.02$ | | | | | $F(5, 266) = 2.05, p = 0.072, R^2 = 0.04$ | | | | |
| PANSS<br>(Wallwork) | Intercept | 0.02 | 0.16 | 0.15 | 0.878 | --- | -0.14 | 0.17 | -0.83 | 0.404 | --- |
|  | Positive | 0.06 | 0.05 | 1.23 | 0.219 | 0.01 | 0.12 | 0.06 | 2.13 | 0.034* | 0.02 |
|  | Negative | 0.03 | 0.05 | 0.77 | 0.441 | 0.00 | 0.04 | 0.05 | 0.69 | 0.488 | 0.00 |
|  | Disorganized/Concrete | -0.03 | 0.06 | -0.53 | 0.599 | 0.00 | 0.06 | 0.06 | 0.87 | 0.386 | 0.00 |
|  | Excited | -0.06 | 0.07 | -0.87 | 0.384 | 0.00 | -0.13 | 0.08 | -1.62 | 0.106 | 0.01 |
|  | Depressed | 0.05 | 0.05 | 1.04 | 0.295 | 0.00 | 0.01 | 0.05 | 0.19 | 0.849 | 0.00 |
| | Model statistics | $F(5, 325) = 0.85, p = 0.514, R^2 = 0.01$ | | | | | $F(5, 325) = 2.46, p = 0.033^*, R^2 = 0.04$ | | | | |

\* Significant at $\alpha = 0.05$ .
PANSS – Positive and Negative Syndrome Scale, SANS – Scale for the Assessment of Negative Symptoms, SAPS – Scale for the Assessment of Positive Symptoms

**Table 3.** Spearman correlation coefficients for illness duration and total scores of depressive scales with NT and PCA model decision scores.

|  | NT model |  |  |  | PCA model |  |  |
| --- | --- | --- | --- | --- | --- | --- | --- |
|  | N | r | R<sup>2</sup> | p | r | R<sup>2</sup> | p |
| Illness Duration | 244 | 0.04 | 0.00 | 0.518 | 0.07 | 0.01 | 0.263 |
| CDSS (total) | 71 | 0.17 | 0.03 | 0.148 | 0.09 | 0.01 | 0.462 |
| HDRS-17 (total) | 219 | -0.07 | 0.00 | 0.305 | 0.00 | 0.00 | 0.984 |

CDSS – Calgary Depression Scale for Schizophrenia, HDRS-17 – HDRS-17 – Hamilton Depression Rating Scale

Subsequently, we further explored associations between decision scores and individual items using separate linear regression models for each scale and filtering for medium effects. For SCZ symptoms, there were no effects between items from the PANSS or SANS and model decision scores. However, SAPS item 35 (Inappropriate Affect) was positively related to both NT and PCA model decision scores (NT: *b* = 0.56, *SE* = 0.25, *t* = 2.23, *p* = .029, η² = .07; PCA: *b* = 0.79, *SE* = 0.25, *t* = 3.18, *p* = .002, η² = .13), indicating medium (NT) to medium-large (PCA) effects, with greater inappropriate affect associated with more SCZ-like model decisions. For depressive symptoms, no HDRS-17 items were associated with either model decision scores. However, CDSS item Depression was positively related to NT decision scores (*b* = .58, *SE* = .29, *t*(61) = 2.02, *p* = .048, η² = .06), whereas Observed Depression was negatively related (*b* = −.79, *SE* = .35, *t*(61) = −2.27, *p* = .027, η² = .08), indicating more SCZ-like model decisions with higher self-reported depression but more HC-like decisions with higher observed depression. Full details are reported in Table 2 and 3 and Supplementary Tables S8 to S12.

#### Association with psychotropic medication exposure

Lastly, no significant differences in decision scores were found between drug-naïve and exposed patients for antipsychotics, antidepressants, benzodiazepines or mood stabilizers in all cohorts (all *p* > 0.05) except for antipsychotic medication exposure on the NT model decision scores in the UCLA cohort. However, antipsychotic medication exposure only explained 9% of the variance in the NT model’s decision scores. While not significant, some medications showed medium to large effects on the model decision scores for specific cohorts (see Supplementary Table S13 for full statistical details).

### Individual feature importance clusters of the NT model

Using k-means clustering (k = 2) on individual feature importance profiles derived from the NT-informed classifier, we identified two robust NT-based subgroups (Cluster 1: N = 237; Cluster 2: N = 497). Examining interactions between cluster assignments and diagnoses, there was a significant association between cluster membership and diagnostic label (*χ²*(1,734) = 7.74, *p* = 0.005). Cluster 2 contained a higher proportion of patients (61.4%) than Cluster 1 (50.2%; see Supplementary Table S15 for diagnostic group percentages). Overall, the two clusters exhibited distinct neurochemical profiles (i.e., filtered for statistical significance and medium effect sizes) with respect to how specific features influenced their classification as SCZ (see Figure 3). In Cluster 1, SCZ predictions were primarily driven by subcortical contributions from COX-1, D2, DAT, and mGluR5. These same features showed the opposite pattern in Cluster 2, generally contributing toward HC classification, with the exception of COX-1, which had minimal influence. In contrast, Cluster 2 showed SCZ-driving contributions from a distinct set of features, including cortical 5-HT4, A4B2, D2, FDOPA, and NMDA, all of which were largely irrelevant for the classification results in Cluster 1. Additionally, Cluster 2 exhibited strong SCZ-driving effects from a broad range of subcortical features, particularly serotonergic (5-HT1A, 5-HT4, 5-HT6), GABAergic (GABAA, GABAA5), as well as HDAC, M1, SV2A, translocator protein (TSPO), and VAChT. These features were either negligible or contributed to the opposite (i.e., HC-driving) direction in Cluster 1. Corresponding statistics, p-values and effect sizes are reported in Supplementary Table S16.

**Figure 3.**
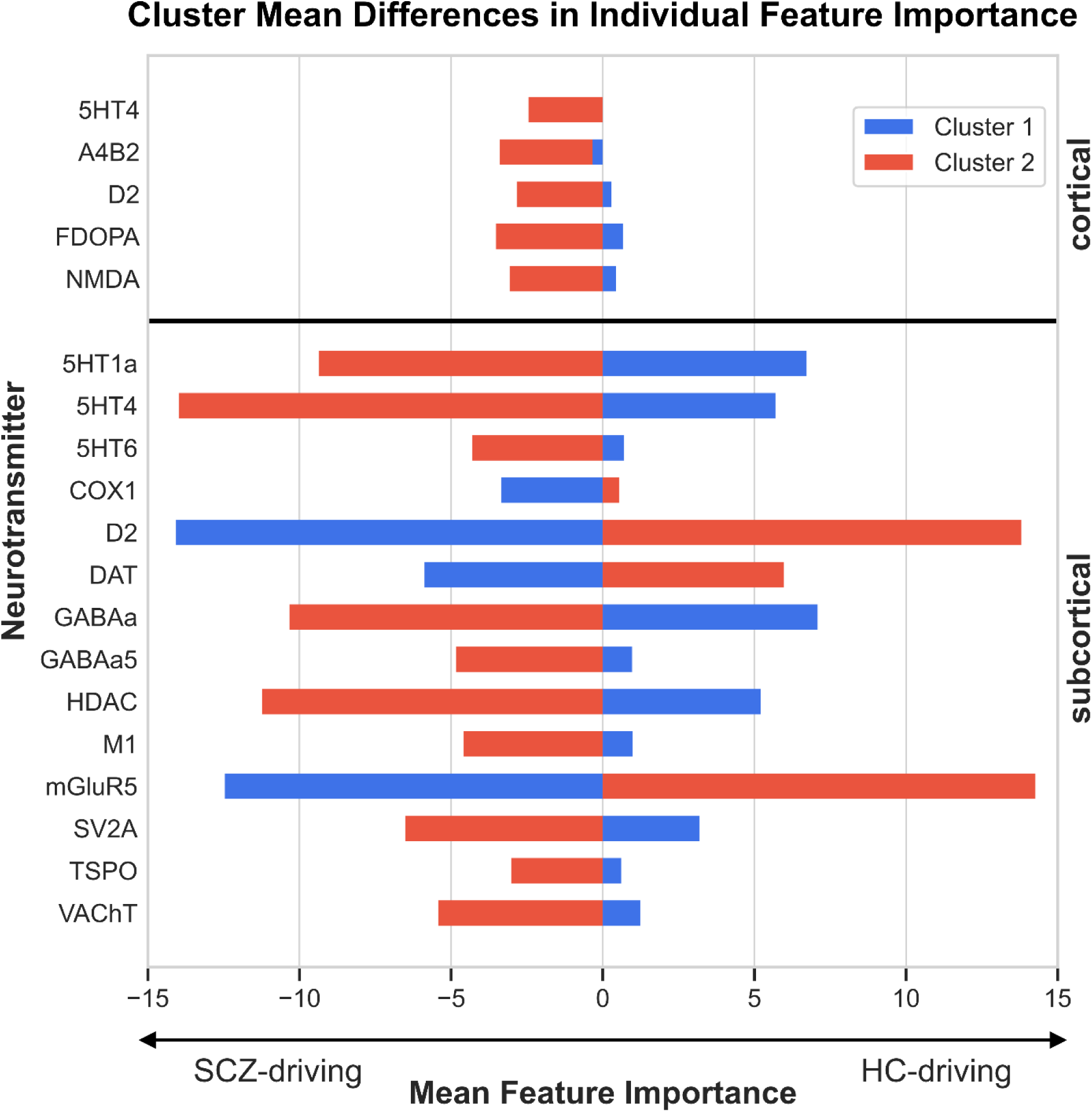
Cluster mean differences in individual feature contributions of the NT model. Cluster mean differences in individual feature contributions (Cluster 1: N = 237; Cluster 2: N = 497) were FDR-corrected for multiple comparisons and filtered for medium effect sizes (|r_z_| ≥ 0.3).

Comparing cluster compactness as measured by distance to centroid, there was a significant difference between clusters (*W* = 68429, *z* = 3.55, *p* = <.001), with Cluster 1 having significantly higher distances to the centroid compared to Cluster 2. When examining the relationship between distance to centroid and symptom burden, there was a significant negative association for Alogia (SANS global rating) in Cluster 1 (*b* = −6.42, *SE* = 3.07, *t*(34) = −2.09, *p* = .044, η² = .12), indicating more severe alogia symptoms closer to the centroid. In Cluster 2, Alogia (SANS global) was also negatively associated with distance to centroid (*b* = -2.59, *SE* = 1.18, *t*(110) = -2.19, *p* = .0.031, η² = .02), though only showing a small effect compared to Cluster 1. Moreover, HDRS-17 total score was negatively correlated with distance to centroid in both clusters (Cluster 1: *r* = −.27, *p* = .143, η² = .07; Cluster 2: *r* = −.27, *p* = .017, η² = .07), indicating more severe depression was associated with a smaller distance to centroid, though this effect was only statistically significant in Cluster 2. Full statistical details are reported in Supplementary Tables S17 and S18.

To further investigate phenotypical differences between clusters, we compared BrainAGE, demographics, and clinical measures. Cluster 1 had significantly higher BrainAGE values than Cluster 2 (*W* = 92953, *z* = 2.18, *p* = 0.029, *r_z_* = 0.08). In contrast, there were no significant differences between clusters regarding age (*W* = 88528, *z* = 0.58, *p* = 0.563) or sex (*χ²(*1) = 1.73, *p* = 0.189). Moreover, comparisons of clinical symptom scores revealed no significant differences between clusters regarding illness duration, SCZ symptom burden (Wallwork PANSS factors, SAPS and SANS global ratings), or depressive symptom burden (total scores of CDSS and HDRS-17) after FDR correction (all *p_FDR_* > 0.05; see Supplementary Table S19).

## Discussion

This study examined whether GMV alterations in SCZ co-localize with normative distributions of NT density maps, and whether these patterns support diagnostic classification (SCZ vs. HC), transdiagnostic application (MDD, BD, ADHD, and BPD), and neurochemically informed subtyping. The NT classifier was able to distinguish individuals with SCZ from HC with a higher sensitivity of 61.8% and a lower specificity of 62.3% compared to the PCA-based model (sensitivity = 58.4%, specificity = 81.9%). For the NT model, SCZ classification was associated exclusively with NT correlations in subcortical regions, showing spatial overlap with serotonergic (5-HT4), endocannabinoid (CB1), and cholinergic (VAChT) NT systems, as well as with markers of synaptic density (SV2A) and gene expression regulation (HDAC). For the PCA model, SCZ classification was mainly based on subcortical and medial temporal structures. Both models primarily showed relatively high diagnostic specificity for SCZ, with only modest transdiagnostic alignment to other psychiatric disorders including BPD and MDD for the NT model, and MDD and BD for the PCA model. Moreover, PCA scores were associated with higher BrainAGE and positive symptoms, particularly hallucinations, leading to more SCZ-like model decisions, whereas NT scores were unrelated to BrainAGE. NT-based clustering revealed two distinct subgroups – one driven by subcortical dopaminergic, glutamatergic, and inflammatory markers, and another involving broader cortical and subcortical serotonergic, dopaminergic, GABAergic, and cholinergic contributions. Stronger alignment with either cluster was linked to higher Alogia, while Cluster 2 also showed higher depressive symptom burden with stronger cluster alignment. Finally, Cluster 1 had higher BrainAGE scores than Cluster 2, with no accompanying differences in clinical symptoms.

Our findings align with a previous report on the relationship between disease-related brain abnormalities and normative NT maps in SCZ. The study also reported positive correlations with D2 and DAT, and negative correlations with 5-HT4 and CB1 ^32^, but without distinguishing between cortical and subcortical regions. Beyond case-control classification, clustering of patients based on individual NT feature importance uncovered two distinct neurochemical subtypes. Cluster 1 is marked by a subcortical inflammatory-dopaminergic-glutamatergic profile, pointing to a mechanism centered on neuroinflammatory and dopaminergic dysregulation within subcortical circuits. Alterations in COX-1, a key enzyme in prostaglandin-mediated neuroinflammation, have been observed in SCZ, including reduced cortical expression and elevated peripheral levels, suggesting a role for inflammatory imbalance in the disorder ^47,48^. The strong contribution of D2 aligns with long-standing evidence for increased striatal D2 receptor availability and heightened subcortical dopaminergic signaling ^49^, whereas DAT findings remain inconsistent, showing no clear increase in transporter availability ^50^. mGluR5 involvement may suggest that glutamatergic dysfunction acts in concert with dopaminergic abnormalities. Although reductions in mGluR5 binding are not consistently observed after controlling for smoking, individual variability in mGluR5 has been linked to negative symptoms and cognitive deficits ^51,52^. Overall, Cluster 1 points to a subcortical subtype involving inflammation-related dopaminergic-glutamatergic imbalance consistent with classical pathophysiological models of SCZ. Compared with the classification framework, in which cortical COX-1 and subcortical D2 and DAT correlations were more strongly associated with HC classification, a shift of these same NT features being SCZ-driving in Cluster 1 indicates that mechanisms linked to preserved neural structure in healthy individuals may become dysregulated, leading to maladaptive rather than compensatory neurochemical-structural relationships.

In contrast, Cluster 2 is characterized by broad cortical and subcortical associations spanning serotonergic, dopaminergic, GABAergic, and cholinergic signaling, alongside markers of gene expression regulation, synaptic density, and neuroimmune activity. Together, these features indicate a distributed neuromodulatory imbalance that disrupts cortical-subcortical integration. The involvement of multiple serotonergic receptors (5-HT1A, 5-HT4, 5-HT6) together with cortical dopaminergic and glutamatergic contributions supports the notion that serotonin-driven modulation of dopamine and glutamate signaling may compensate for or exacerbate broader network dysregulation ^8,13,53–59^. The accompanying GABAergic and cholinergic associations further point to disrupted excitatory-inhibitory balance, consistent with cortical disinhibition models of SCZ ^60–63^. Additional associations with SV2A and HDAC markers indicate that these widespread neurochemical effects may extend to synaptic integrity and transcriptional control, implying a system-level disturbance in synaptic plasticity and gene expression ^64–66^. GMV reductions may partly index synaptic loss, as GM decreases have been linked to reductions in dendritic and spine density ^67,68^. This fits with the synaptic hypothesis of SCZ, which proposes that primary disturbances in synaptic integrity drive downstream NT dysregulation and large-scale circuit abnormalities ^69^, offering a mechanistic bridge between SV2A-related findings and the NT-system alterations observed here. Furthermore, the findings aligns with proposals that HDAC-mediated chromatin remodeling contributes to SCZ pathophysiology and highlights HDAC inhibitors as potential targets for epigenetic intervention ^70^. The presence of TSPO-related neuroimmune markers suggests that low-grade neuroinflammatory activity could contribute to this distributed pattern ^71^. Altogether, Cluster 2 appears to represent a diffuse, multi-system mechanism characterized by interdependent serotonergic, GABAergic, and cholinergic dysregulation, consistent with models emphasizing impaired cortical coordination and neuromodulatory homeostasis in SCZ ^56,60–62,72^. Compared with the classification pattern, Cluster 2 aligns strongly with features related to SCZ classification including 5-HT4, HDAC, SV2A, and VAChT. While neither cluster included CB1, which was relevant for SCZ classification in the NT model, Cluster 2 broadens into a cortical-subcortical serotonergic-GABAergic-cholinergic disturbance.

Conceptually, these two NT-based clusters align with the structural subtypes reported by Dwyer and colleagues ^73^ – one characterized by rather circumscribed cortical volume reductions, the other showing more diffuse cortical and subcortical volume reductions – suggesting convergent evidence for at least two mechanistically distinct profiles underlying SCZ. While the current study also found evidence for a mechanistically diffuse and a more specific subtype, the latter was restricted to subcortical correlations. This discrepancy is plausibly attributable to our transdiagnostic sample, as well as the fact that clustering was performed on model-derived attributions. Given that cluster labels improved accuracy compared to case-control labels, examining models trained on NT-correlation-derived cluster labels is warranted. Taken together, the NT-informed model highlights mechanistically relevant heterogeneity in SCZ, revealing system-level variation that reflects the underlying biological complexity of the disorder. Moreover, our findings highlight the potential of a normative NT mapping approach to reveal neurochemical correlates of SCZ-related brain changes that may be overlooked in traditional PET studies, which remain limited by small sample sizes. Such NT-informed stratification could help identify biologically defined subgroups that differ in their responsiveness to specific pharmacological targets, thereby informing more personalized treatment approaches.

The performance of the PCA-based model is in line with prior GMV-based SCZ classifiers, which typically report BAC values ranging from 65% to 75% ^39,74–76^. Regionally, SCZ decisions were driven mainly by subcortical structures (thalamus, hippocampus) commonly showing GMV loss, whereas HC decisions relied on distributed cortical regions (frontal, temporal, parietal, occipital) ^19,39^. This pattern aligns with prior work indicating that both cortical and subcortical abnormalities are central to SCZ, but their relative impact on classification depends on the model and input features ^39,76^. The lower specificity of the NT model may reflect less clearly defined NT-GMV patterns in HC, while more systematic deviations in SCZ enable the model to identify patients with comparatively good sensitivity. The moderate correlation between model decisions indicates that the NT model is not merely replicating the PCA model’s predictions. However, the poor performance of the stacked model suggests these signals are not sufficiently complementary to boost classification. Consistent with prior work, PCA model decision scores were linked to accelerated brain aging across diagnostic groups ^23^, whereas NT scores were not, implying that NT-related structural deviations may occur independently of normative age trajectories.

Comparing the transdiagnostic generalizability of the classifiers, the NT model had modest success for the classification of BPD and MDD, suggesting transdiagnostic overlap. This aligns with evidence of serotonergic, and dopaminergic dysfunction in BPD and MDD ^77–79^, but also indicates that the NT-informed pattern may be more specific to SCZ. The lower sensitivity compared to specificity in these disorders suggests limited detection of subtle or heterogeneous structural alterations across conditions. Similarly, the PCA model also displayed limited generalizability to MDD and additionally BD, which is in line with BD showing substantial overlap with SCZ in terms of risk genes and symptoms ^80^.

Overall, only few associations between classifier decision scores and symptom burden were found, reinforcing longstanding challenges in mapping brain structure to clinical features in SCZ ^81^. Similarly, the mechanistically defined subtypes did not differ by age, sex, illness duration, or overall symptom burden. However, Cluster 1 exhibited significantly higher BrainAGE, consistent with the hypothesis that accelerated brain aging may reflect more severe or biologically active forms of the illness ^82^. Distance-to-centroid analyses further indicated that alogia and depressive symptoms were linked to increased cluster alignment. The absence of pronounced clinical differences between clusters aligns with the view that SCZ reflects multiple mechanistic pathways converging on similar phenotypic outcomes ^83,84^. Moreover, given the transdiagnostic basis of the clustering, identifying a single shared clinical phenotype may be inherently difficult. Overall, NT-based clustering may reveal biologically distinct subgroups within SCZ, even in the absence of clear clinical stratification, underscoring the need for larger and more phenotypically rich datasets to validate and further characterize these neurochemical subtypes.

Several limitations need to be mentioned regarding our findings. First, psychotropic medication use, which can influence both NT receptor availability and brain structure ^85,86^, could not be fully accounted for due to missing or non-harmonized medication data across cohorts. Notably, medication information was available for only a small subsample of the MUC cohort. Future studies should test whether these patterns replicate in drug-naïve patients and in earlier disease stages (e.g., at clinical high risk for psychosis). Second, the normative NT density maps were derived from independent healthy volunteer samples, introducing the possibility of spatial mismatches or limited applicability to clinical populations, though the availability of PET or SPECT studies is limited and commonly includes small sample sizes. Third, the directionality analysis for the NT correlations revealed widespread correlations in patients, limiting the interpretability of the mean directionality for each NT. However, this further stresses the need for mechanistically and clinically meaningful subgroups in SCZ. Future studies should verify and refine this stratification by acquiring PET/SPECT data in patient subgroups (e.g., Cluster 1 vs. Cluster 2) to test the predicted NT-system differences and directional patterns. Fourth, the modest classification accuracy of NT-based models, while statistically significant, remains insufficient for clinical implementation. Future studies should revisit this question using stratified, mechanistically informed subgroups rather than a simple case-control framework.

Finally, associations with clinical symptoms were limited by inconsistent scale coverage across cohorts, small subgroup sizes, and missing data, which together constrain generalizability. Additional incorporation of direct PET imaging of receptor availability in patients would allow validation of NT co-localization hypotheses. Finally, exploring whether these NT-informed subtypes differ in response to pharmacological interventions may help guide mechanism-based treatment stratification.

Our findings suggest that SCZ-related structural brain changes exhibit spatial overlap with normative NT distributions, revealing two mechanistic subtypes in SCZ with differential neurochemical NT system-specific vulnerabilities. Although less specific than GMV-only models, this approach offered greater sensitivity and biological interpretability, helping to explain why case-control analyses miss key heterogeneity. These insights may contribute to more personalized and biologically grounded approaches in psychiatric research and treatment.

## Acknowledgement

COBRE data were downloaded from the COllaborative Informatics and Neuroimaging Suite Data Exchange tool (COINS; http://coins.mrn.org/dx) and data collection was performed at the Mind Research Network, and funded by a Center of Biomedical Research Excellence (COBRE) grant 5P20RR021938/P20GM103472 from the NIH to Dr. Vince Calhoun.

The MCIC imaging data and demographic information was collected and shared by [University of Iowa, University of Minnesota, University of New Mexico, Massachusetts General Hospital] the Mind Research Network supported by the Department of Energy under Award Number DE-FG02-08ER64581.

## Data Availability

The original MUC and MIMICSS data and their derivatives cannot be made publicly available as the study includes sensitive patient data and public data sharing was not covered in the informed consent. The original data supporting the findings of this study are available from Florian Raabe (MIMICSS) or Nikolaos Koutsouleris (MUC) upon reasonable request. The COBRE, MCIC, and UCLA cohorts are publicly available through the SchizConnect platform at https://schizconnect.org/.

## Author contributions

LH conceptualized the project, conducted the analyses, and drafted the manuscript. NK contributed to the conceptualization and was involved in data collection. JF supported data processing. CV contributed to the implementation of the methodology in NeuroMiner. FR, DK, AH, IP and EM were involved in data collection. All authors critically revised the manuscript and approved the final version for publication.

