## Supplement for "Mapping Neurochemical Signatures onto Brain Structure for Neurotransmitter-Informed Discrimination of Schizophrenia Patients from Healthy Controls"

### Supplementary Information

#### Supplementary Methods

##### MRI data acquisition, preprocessing and covariate correction

Within the COBRE and MCIC cohorts, the participants had multiple scans per visit including either multiple runs (COBRE and MCIC), different echo gradients (CORBE only), or a combination of both (COBRE only). In the case of availability of multiple scans per visit, mean T1-weighted images were calculated from co-registered scans (see Supplementary Table S3 for the number of available scans per cohort). Images were not smoothed, as the computation of neurotransmitter (NT) correlations relies on unsmoothed data.

Prior to analyses, all images were corrected for age, sex and site (i.e., scanner) effects combining a mean centering approach with a dynamic standardization procedure (1). First, site-specific effects among healthy controls (HC) were estimated by computing voxel-wise differences between 23 individual site-based HC groups (total N = 184; mean age = 31.8 ± 9.3 years; 95 females), each matched for age and sex across sites, and the overall mean gray matter volume (GMV) map derived from the entire HC sample. This mean-centering approach was then applied to all images to remove site-related variability from the GMV maps. Next, we applied a dynamic standardization procedure to adjust each participant's GMV map for normative age- and sex-related changes. For each individual, we constructed a normative reference sample from the offset-corrected HC data (N = 414) by selecting participants within an age range of ± 6 years and of the same sex. In the case of HC, the normative sample excluded the participant currently being standardized. Finally, voxel-wise medians and standard deviations were calculated from the normative sample and used to standardize the participant's GMV map accordingly.

##### Support vector classification

We constructed and tested two unimodal classifiers, an NT-informed classifier and a mechanism-agnostic, purely structural-imaging-based classifier, to differentiate SCZ from HC to evaluate differences in accuracy and in the phenotypic profile identified by each model. Using a nested outer leave-one-site-out cross-validation (LOSOCV) framework including ten repetitions of eight outer CV (CV2) folds and one repetition of ten inner CV (CV1) folds, classification was based on covariate-corrected GMV maps. To avoid information leakage between training and test samples, preprocessing parameters were computed within the CV1 training sample of the nested CV design. For the NT model, partial Spearman correlation coefficients were calculated between cortical and subcortical GMV and the normative distribution of NT density maps using the JuSpace methodology (see Supplementary Methods for details about the computation of the NT correlations). For the second model, this preprocessing step was replaced by a principal component analysis (PCA) for dimensionality reduction, which was restricted to 50 principal components to match the number of features of the NT model. Here, the PCA model served as a control model to enable the comparison of classification performance and identification of phenotypic profile. The resulting features of each model pipeline were then standardized and forwarded to a greedy forward search wrapper algorithm involving the linear sequential minimal optimization C-support vector machine (C-SVM; LIBSVM version 3.12, LIBSVM library (2)). Performance was evaluated using balanced accuracy (BAC; i.e., mean of sensitivity and specificity). Additionally, a stacked model was implemented to assess whether combining predictions from the NT and PCA models could improve classification performance. Finally, Quade tests (3) were employed to evaluate differences in the models' median BAC, calculated from the CV2 test data partitions (10 permutations of 8-fold CV2).

To assess similarity of the decision scores between both models, a Spearman correlation coefficient and the corresponding $R^2$ was calculated between the decision scores. To assess the statistical significance of the overall model performance (i.e., extent to which the result may be replicated), label permutations were performed. In this process, class labels were randomly shuffled multiple times, and the model was retrained on each permuted dataset to generate a null distribution of performance metrics. The model's actual performance was then compared against this null distribution to determine whether it exceeded chance levels at $\alpha = 0.05$. This method is described in detail in the supplementary material of Koutsouleris et al. (2016) (4).

#### Computation of correlations with the normative distribution of neurotransmitter density maps

To compute correlations between sample-dependent GMV images and the normative distribution of NT density maps (i.e., maps derived from a healthy volunteer population using PET or SPECT), the methodology of the JuSpace toolbox (5) was integrated into our in-house machine learning software NeuroMiner version 1.4 (https://github.com/neurominer-git/NeuroMiner) (6). More specifically, spatial correlations were computed between covariate-adjusted GMV images and nuclear imaging-derived maps containing information about the relative density distribution of NT systems in independent HC populations. More specifically, mean values were extracted from NT and GMV maps using the Schaefer atlas containing 100 cortical regions and the Tian atlas containing 54 subcortical regions. Fisher's z-transformed Spearman correlation coefficients were then calculated between z-transformed GMV maps and 25 NT maps for cortical and subcortical atlases separately. Partial correlation coefficients were used to adjust for spatial autocorrelation and partial volume using the tissue probability map provided by SPM12 (Wellcome Trust Centre for Neuroimaging, London, UK; www.fil.ion.ucl.ac.uk/spm). Details about the utilized NT maps including information regarding tracers and HC populations are reported in Supplementary Table S4.

#### Directional Inference via Cross-Validated Neurotransmitter Correlation

To facilitate the biological interpretability of the ML-based classification results, particularly the directionality of imaging alterations, we conducted a supplementary analysis computing correlations between z-scored GMV values and normative NT density maps. For each outer CV2 permutation and fold, we extracted region-wise GMV values from patients and matched HC using the Schaefer (100 cortical regions) and Tian (54 subcortical regions) atlases. GMV values from patients were z-transformed based on the mean and standard deviation of controls within each fold, ensuring that the resulting measures reflected deviation from normative brain structure. Spatial Spearman correlations were then computed between these z-transformed GMV maps and 25 normative NT density maps separately for cortical and subcortical regions. This approach allows the interpretation of whether increases or decreases in GMV of patients relative to HC co-localize with specific NT systems. Correlation coefficients were Fisher z-transformed and averaged across folds and permutations for each individual to obtain stable subject-level estimates. Final group-level results were summarized by computing the mean and standard deviation of these subject-level correlation values for each NT map.

#### Computation of BrainAGE

BrainAGE was computed to examine the impact on the decision scores (7). A linear ν-support vector regression (ν-SVR) was used to predict age from each individual's GMV map by training the predictive model in all 847 HC (mean age = 33.3 ± 11.1 years; 368 females) within the same nested CV design (i.e., LOSOCV with 10x8 for CV2 and 1x10 for CV1) using the mean absolute error (MAE) as the optimization criterion. The preprocessing pipeline included smoothing using a

Gaussian convolution kernel (i.e., 4 mm, 6 mm, 8 mm, and 10 mm) full-width at half maximum, adjustment for sex effects using partial correlation, correction for site effects using global mean correction, dimensionality reduction via PCA (retaining components explaining 80% of the variance), standardization of the resulting principal components with values limited to ±4 standard deviations, and voxel-wise scaling wrapped within each training cycle. The beta coefficients for partial correlations and the global means were computed in the 184 age- and sex-matched HC (mean age = 31.8 ± 9.3 years; 95 females) and then applied to all HC. The normative model was subsequently applied across all patient groups. Similar to the classification models, CVR mapping and a sign-based consistency approach were used to evaluate stability of predictive pattern components and the importance of predictive features. To address the tendency for age predictions to be overestimated in younger individuals and underestimated in older individuals, a linear regression-based correction was applied to the predicted age values (8). Beta coefficients were estimated in the HC sample using partial correlations and then applied to the patient sample. Finally, BrainAGE scores were calculated for all groups by subtracting chronological age from the tail-corrected predicted age.

#### Availability of clinical information including illness duration, psychotic and depressive symptoms, and medication information

Illness duration was computed for the MIMICSS dataset as difference between age at the point of study inclusion and age of onset and for the COBRE dataset as difference between at the point of study inclusion and age at first psychotic symptoms. For the MCIC dataset, the information was readily available, but not for MUC and UCLA cohorts.

Regarding psychotic symptoms, for COBRE, MIMICSS and UCLA cohorts, the Positive and Negative Syndrome Scale (PANSS) (9) was available, whereas the Scale for the Assessment of Negative Symptoms (SANS) (10) was available for COBRE, MIMICSS, and UCLA cohorts. The Scale for the Assessment of Positive Symptoms (SAPS) (11) was only available for MCIC and UCLA cohorts. For PANSS, Spearman correlation coefficients were computed for positive, negative and general scores in addition to the total score. For SAPS and SANS, Spearman correlation coefficients were computed for sum scores. Correlation coefficients were computed for each patient group separately (schizophrenia [SCZ]: PANSS, SAPS, SANS; bipolar disorder [BD]: SANS, SAPS; clinical high risk for psychosis CHR-P]: PANSS; borderline personality disorder [BPD]: PANSS) and p-values were FDR-corrected for the number of correlations with total scores within each patient group.

Regarding depressive symptoms, the Calgary Depression Scale for Schizophrenia (CDSS) (12) was available for COBRE, and the Hamilton Depression Rating Scale (HDRS-17) (13) was available for both MUC and UCLA. For all depression scales, Spearman correlation coefficients were computed for total scores of each scale and for patient group separately (SCZ: CDSS, HDRS-17; major depressive disorder [MDD]: HDRS-17; BD: HDRS-17; attention deficit hyperactivity disorder [ADHD]: HDRS-17). Corresponding p-values were FDR-corrected for the number of correlations with total scores within each patient group.

Medication data were available for patients in the UCLA, COBRE, MIMICSS, and MCIC (i.e. neuroleptic naïve status) cohorts including SCZ, BD and ADHD patients. In the MUC cohort, medication information was available for only 24 patients, all of whom had been exposed to antipsychotics, precluding a drug-naïve vs. exposed comparison. For MUC, we therefore replaced the binary exposure indicator with chlorpromazine (CPZ)-equivalent dose. Exposure was characterized as current or prior use of a medication within the relevant category.

#### Computation and clustering of individual feature importance

To quantify the contribution of individual features to the predictions of the NT model, we utilized the median flip method, a perturbation-based feature importance approach integrated into the ML software NeuroMiner (6). Specifically, each feature's percentile value was flipped across the median by shifting it by 50 percentile points (i.e., values below the 50th percentile were increased by 50 and values above the 50th percentile were decreased by 50), while all other features remained unchanged. The change in the model's prediction following this inversion was used to compute a feature importance score, summarized as the median change. To obtain individualized feature importance profiles for the NT model, we replaced each individual's original imaging data (i.e., covariate-corrected voxel-wise GMV images) with their CV1-average of the preprocessed NT correlations, ensuring that the median flip perturbations were applied directly to the NT correlation features.

To reveal potential mechanistic subtypes based on individual feature importance, we performed clustering using data-driven, unsupervised machine learning techniques on the feature importance scores of the individuals classified as patients, following a similar approach to Oeztuerk et al. (2022) (14). Initially, we evaluated four clustering algorithms including k-means, partitioning around meriods (PAM), hierarchical clustering, and agglomerative nesting (agnes), considering both average and Ward linkage methods for hierarchical clustering and agnes. Using the ClValid package (15), cluster solutions ranging from 2 to 10 clusters were considered. To identify the optimal algorithm and number of clusters, we applied a majority rule across multiple internal validity and stability metrics (see Supplementary Table S14). As a result, hierarchical clustering with average linkage and k-means were identified as top-performing algorithms, with k = 2 as the optimal number of clusters. Next, we re-evaluated the top-performing clustering algorithms using the NbClust package (16), which recommends the optimal number of clusters based on a majority rule across 26 internal validity indices (e.g., Calinski-Harabasz, Davies-Bouldin, Silhouette). For k-means clustering, a two-cluster solution was most frequently recommended (13 of 26 indices), whereas for hierarchical clustering with average linkage, two- and five-cluster solutions were most frequently recommended (9 of 26 indices each).

To assess the robustness of these clusters, we used the clusterboot function from the *fpc* package (17) performing 500 resamples to calculate Jaccard similarity for k-means and hierarchical clustering (average linkage) using k = 2 for both methods and k = 5 exclusively for hierarchical clustering. k-means with k = 2 demonstrated the highest overall stability, with mean Jaccard indices exceeding 0.98 (hierarchical with average linkage and k = 2: 0.998 [cluster 1] 0.556 [cluster 2]; hierarchical with average linkage and k = 5: 0.994 [cluster 1] 0.876 [cluster 2] 0.592 [cluster 3] 0.588 [cluster 4] 0.612 [cluster 5];k-means with k = 2: 0. 991 [cluster 1], 0.983 [cluster 2]). To further validate the reliability of the k-means two-cluster solution, we applied the prediction.strength function from the *fpc* package (17) using 500 resamples, which supported k = 2 as a reliable choice (prediction strengths: 1.00 [cluster 1], 0.85 [cluster 2]). Next, clusters were assigned using k-means with k = 2, resulting in 237 as patient classified subjects in cluster 1 and 497 as patient classified subjects in cluster 2.

### Supplementary Results

#### Statistical comparison of model performance and stacked model

Comparing the performance between models, the PCA model performed significantly better than the NT model ($W = 3240$, $z = 7.77$, $p < 0.001$). The stacked model performed significantly worse than the PCA model with a BAC of 69.5% (sensitivity = 57.8%, specificity = 81.2%; $W = -1213$, $z = 3.62$, $p < 0.001$), but significantly better than the NT model ($W = 3202$, $z = 7.68$, $p < 0.0001$).

#### BrainAGE model performance and relevant features

The BrainAGE model performed with a MAE of 5.5 years in HC ($r$ = 0.79, $R^2$ = 61.6; Figure 2A) and a MAE of 6.6 years in patient groups ($r$ = 0.82, $R^2$ = 66.6; SCZ: MAE = 6.7; MDD: MAE = 6.6; BD: MAE = 6.2; CHR-P: MAE = 5.9; ADHD: MAE = 6.3; BPD: MAE = 7.6; see Supplementary Figure S3A). At a stability threshold (CVR) of ≥ |2| and masked by significant sign-based consistencies ($p_{FDR}$ < 0.05), the resulting CVR maps revealed stable features predicting age in HC. There were negative GMV associations predominantly in frontal, temporal, parietal, and occipital cortices, specifically within the superior and middle frontal gyri, superior parietal cortex, and superior and middle temporal gyri. In contrast, positive GMV associations were present in subcortical regions, particularly involving the basal ganglia structures such as the putamen and caudate nucleus, as well as portions of the thalamus and medial temporal areas (see Supplementary Figure S3B).

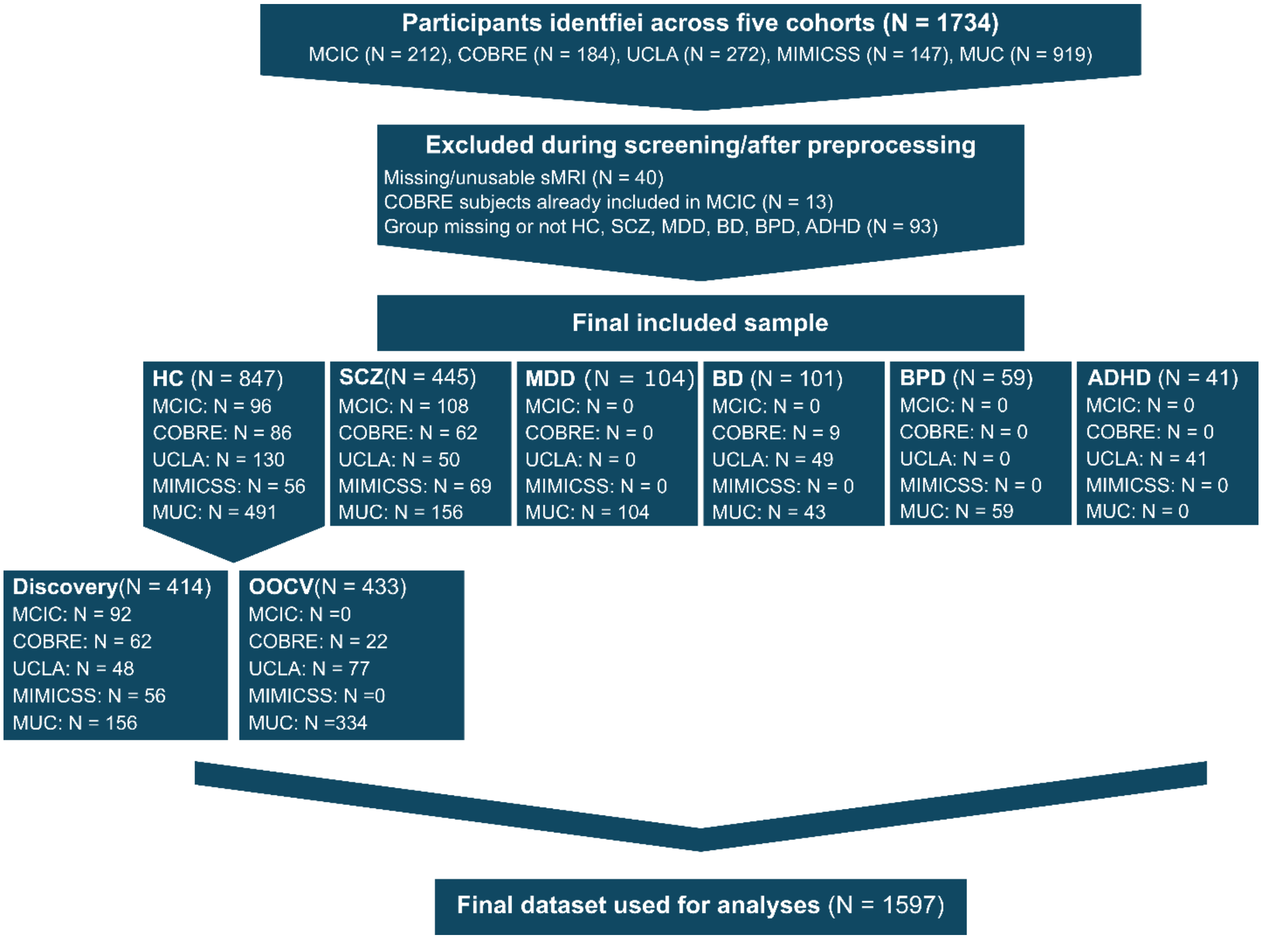


**Figure S1. Flowchart of participant inclusion, exclusion, and final sample composition across the five cohorts.**

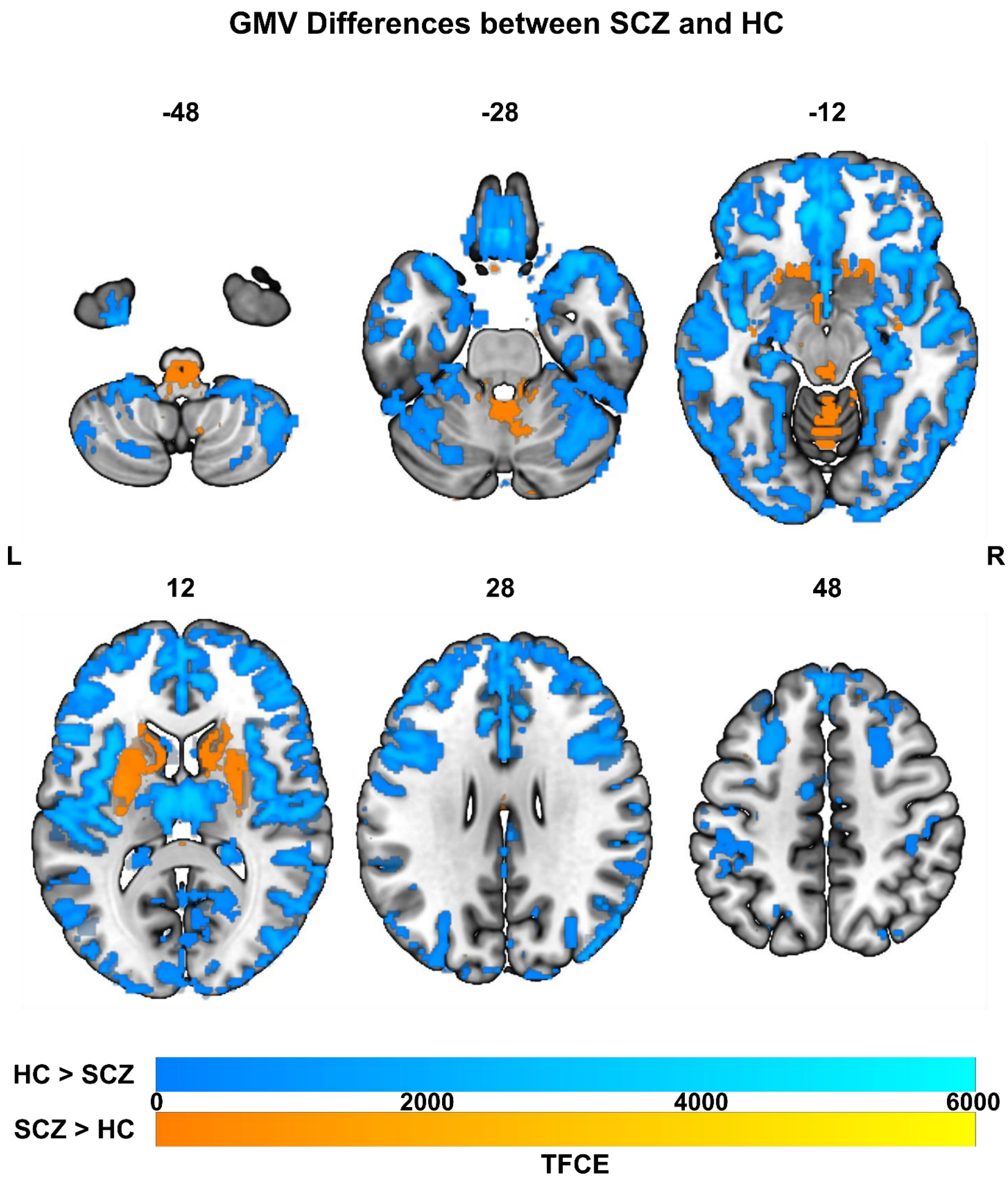


**Figure S2. GMV differences between SCZ and HC.** FDR-corrected (α = 0.05) TFCE contrast map for HC > SCZ (blue) and SCZ > HC (orange).

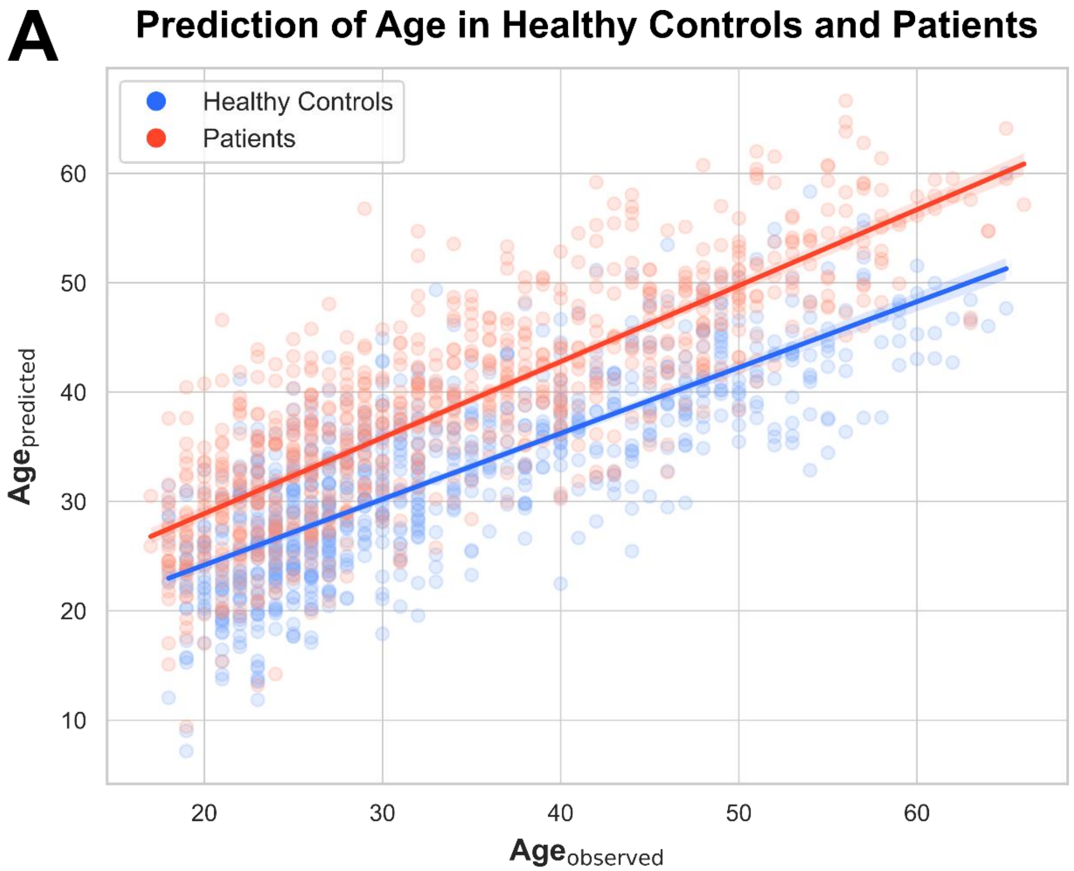
A
Prediction of Age in Healthy Controls and Patients
Healthy Controls
Patients
60
50
40
30
20
10
Agepredicted
20
30
40
50
60
Ageobserved

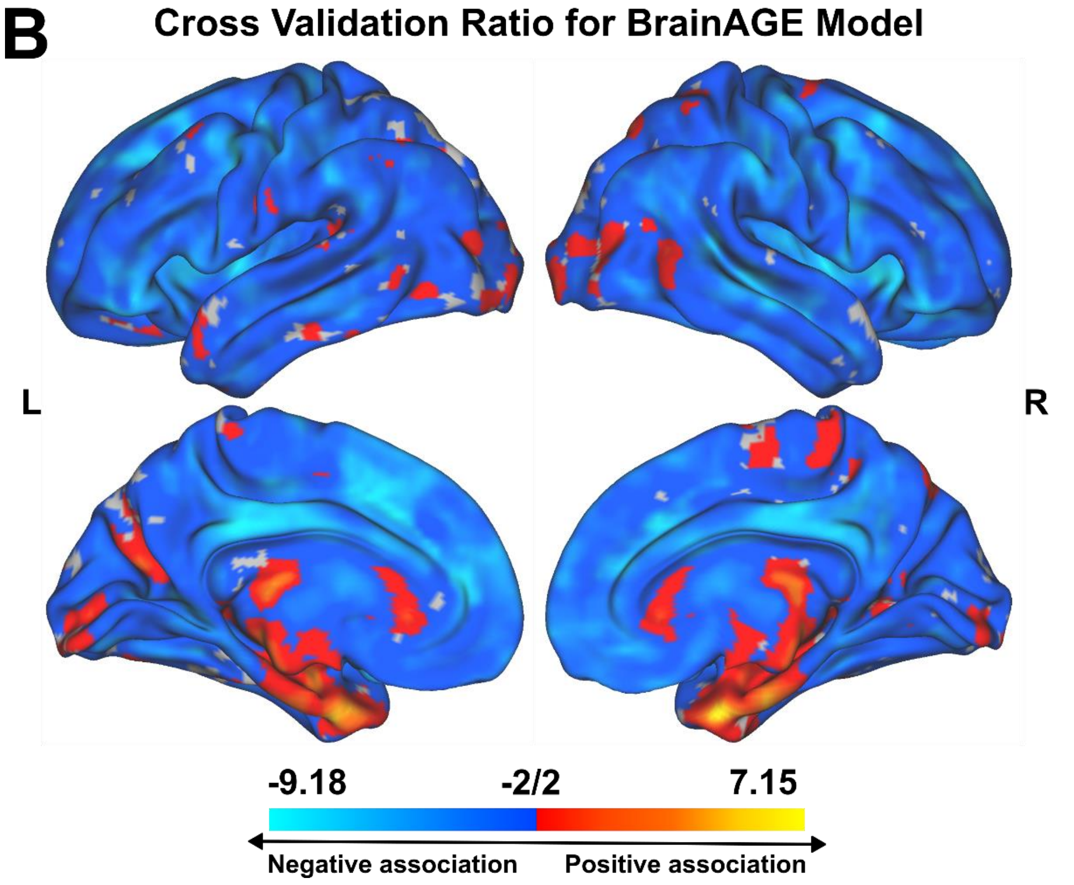
B
Cross Validation Ratio for BrainAGE Model
L
R
-9.18
-2/2
7.15
Negative association
Positive association

**Figure S3. Regression plot and masked cross validation ratio map for the BrainAGE model.** **(A)** Regression plot for the prediction of age in healthy controls and patients. **(B)** **T**he **c**ross-validation ratio map was masked by FDR-adjusted sign-based consistency values for the BrainAGE model.

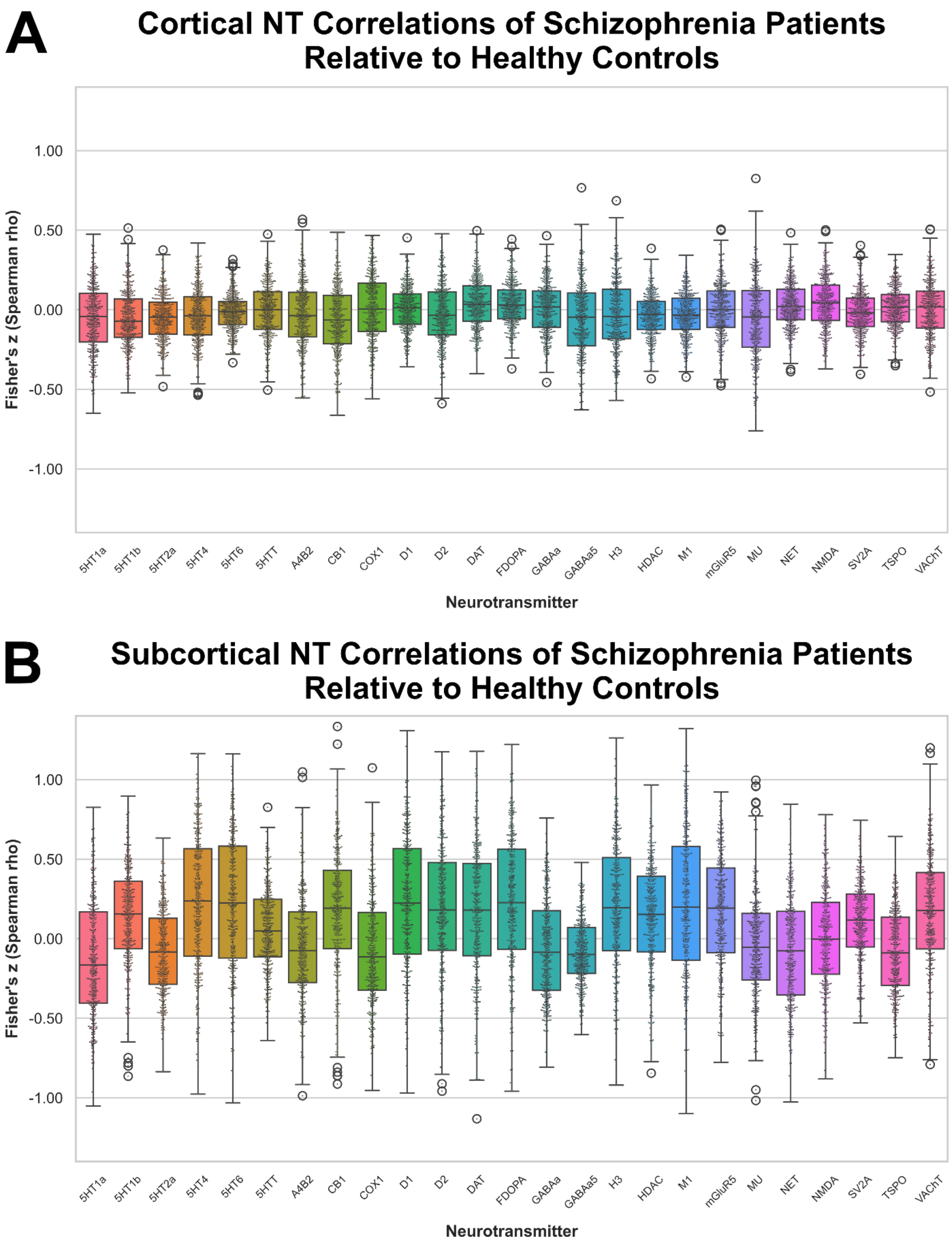


**Figure S4. Cortical and subcortical NT correlations of schizophrenia patients relative to healthy controls.** Directionality analysis of **(A)** cortical and **(B)** subcortical NT correlations of schizophrenia patients relative to healthy controls.

**Table S1.** Demographic and clinical information for HC, SCZ, MDD, BD, CHR-P, ADHD, and BPD across sites within cohorts.

| Group | Cohort | Site | N | Age (years) | Sex (M/F) |
|---|---|---|---|---|---|
| | ALL | - - - | 847 | 33.3 ± 11.1 | 479/368 |
| | COBRE | - - - | 84 | 38.6 ± 11.7 | 62/22 |
| | | A | 43 | 29.0 ± 11.7 | 34/9 |
| | MCIC | C | 24 | 31.9 ± 11.8 | 14/10 |
| HC | | D | 25 | 41.0 ± 9.3 | 15/10 |
| | MIMICSS | - - - | 56 | 33.1 ± 11.8 | 40/16 |
| | MUC | - - - | 490 | 33.0 ± 11.0 | 248/242 |
| | UCLA | A | 102 | 31.7 ± 8.9 | 55/47 |
| | | B | 23 | 30.7 ± 8.6 | 11/12 |
| | ALL | - - - | 414 | 34.3 ± 12.1 | 229/185 |
| | COBRE | - - - | 62 | 35.8 ± 11.4 | 44/18 |
| | | A | 43 | 29.0 ± 11.7 | 34/9 |
| | MCIC | C | 24 | 31.9 ± 11.8 | 14/10 |
| HC (matched)[a] | | D | 25 | 41.0 ± 9.3 | 15/10 |
| | MIMICSS | - - - | 56 | 33.1 ± 11.8 | 40/16 |
| | MUC | - - - | 156 | 35.9 ± 13.2 | 59/97 |
| | UCLA | A | 25 | 31.4 ± 9.1 | 12/13 |
| | | B | 23 | 30.7 ± 8.6 | 11/12 |
| | ALL | - - - | 445 | 34.0 ± 11.3 | 342/103 |
| | COBRE | - - - | 62 | 37.3 ± 13.8 | 51/11 |
| | | A | 44 | 32.8 ± 12.0 | 34/10 |
| | MCIC | C | 31 | 32.2 ± 10.3 | 24/7 |
| SCZ | | D | 33 | 38.3 ± 10.0 | 25/8 |
| | MIMICSS | - - - | 69 | 35.3 ± 11.9 | 54/15 |
| | MUC | - - - | 156 | 31.0 ± 10.1 | 116/40 |
| | UCLA | A | 25 | 36.9 ± 9.3 | 17/8 |
| | | B | 25 | 26.0 ± 8.6 | 21/4 |
| | ALL | - - - | 104 | 42.3 ± 12.0 | 52/52 |
| | COBRE | - - - | | - - - | |
| | | A | | - - - | |
| | MCIC | C | | - - - | |
| MDD | | D | | - - - | |
| | MIMICSS | - - - | | - - - | |
| | MUC | - - - | 104 | 42.3 ± 12.0 | 52/52 |
| | UCLA | A | | - - - | |
| | | B | | - - - | |
| | ALL | - - - | 101 | 39.2 ± 11.0 | 53/48 |
| | COBRE | - - - | 9 | 45.9 ± 15.1 | 5/4 |
| BD | | A | | - - - | |
| | MCIC | C | | - - - | |
| | | D | | - - - | |

| | | | | | |
|---|---|---|---|---|---|
| | MIMICSS | - - - | | - - - | |
| | MUC | - - - | 43 | 42.2 ± 10.8 | 20/23 |
| | UCLA | A | 26 | 34.6 ± 9.5 | 12/14 |
| | | B | 23 | 36.1 ± 8.6 | 16/7 |
| ADHD | ALL | - - - | 41 | 32.3 ± 10.4 | 21/20 |
| | COBRE | - - - | | - - - | |
| | MCIC | A | | - - - | |
| | | C | | - - - | |
| | | D | | - - - | |
| | MIMICSS | - - - | | - - - | |
| | MUC | - - - | | - - - | |
| | UCLA | A | 21 | 33.1 ± 10.4 | 8/13 |
| | | B | 20 | 31.6 ± 10.7 | 13/7 |
| BPD | ALL | - - - | 59 | 25.9 ± 7.0 | 0/59 |
| | COBRE | - - - | | - - - | |
| | MCIC | A | | - - - | |
| | | C | | - - - | |
| | | D | | - - - | |
| | MIMICSS | - - - | | - - - | |
| | MUC | - - - | 59 | 25.9 ± 7.0 | 0/59 |
| | UCLA | A | | - - - | |
| | | B | | - - - | |

[a] Age- and sex-matched (i.e., to SCZ patients) subset of HC.

ADHD – Attention Deficit Hyperactivity Disorder, BD – Bipolar Disorder, BPD – Borderline Personality Disorder, COBRE – Center for Biomedical Research Excellence, MCIC – MIND Clinical Imaging Consortium, MDD – Major Depression, MIMICSS – Multimodal Imaging in Chronic Schizophrenia Study, MUC – Munich Imaging Database, UCLA – UCLA Consortium for Neuropsychiatric Phenomics LA5c Study, SCZ – Schizophrenia

**Table S2.** Site-specific imaging parameters for structural T1-weighted MRI.

| Cohort – Site | Scanner | TE (ms) | TR (ms) | FOV (X, Y, Z) | Voxel size (mm) |
|---|---|---|---|---|---|
| COBRE | Siemens Trio 3T | 1.64<br>3.50<br>5.36<br>7.22<br>9.08 | 2530 | 256 (256 x 256 x 256) | 1 x 1 x 1 |
| MCIC – Site A | Siemens Sonata 1.5 T | 4.76 | 12 | 160 (256 x 256 x 128) | 0.625 × 0.625 x 1.5 |
| MCIC – Site B | GE Signa 1.5 T | 4.76 | 12 | 160 (256 x 256 x 128) | 0.625 × 0.625 x 1.5 |
| MCIC – Site C | Siemens Trio 3T | 3.79 | 2530 | 160 (256 x 256 x 128) | 0.625 × 0.625 x 1.5 |
| MCIC – Site D | Siemens 1.5 T | 4.76 | 12 | 160 (256 x 256 x 128) | 0.625 × 0.625 x 1.5 |
| MIMICSS | Siemens Skyra 3 T | 2.22 | 1900 | 200 (350 x 350 x 263) | 0.8 x 0.8 x 0.8 |
| MUC | Siemens Vision 1.5 T | 4.90 | 11.6 | 230 (512 x 512 x 126) | 0.45 x 0.45 x 1.5 |
| UCLA – Site A/B* | Siemens Trio 3 T | 2.26 | 1.9 | 250 (256 x 256 x 176) | 1 x 1 x 1 |

* UCLA sites differ regarding the scanners' software version.

COBRE – Center for Biomedical Research Excellence, MCIC – MIND Clinical Imaging Consortium, MIMICSS – Multimodal Imaging in Chronic Schizophrenia Study, MUC – Munich Imaging Database, UCLA – UCLA Consortium for Neuropsychiatric Phenomics LA5c Study

**Table S3.** Cohort-specific information about the number of scans and the number of subjects.

| Cohort | Number of scans | Number of subjects |
|---|---|---|
| COBRE | 6 | 112 |
| | 7 | 3 |
| | 12 | 35 |
| | 13 | 2 |
| | 18 | 3 |
| MCIC | 1 | 3 |
| | 2 | 55 |
| | 3 | 120 |
| | 4 | 22 |
| MIMICS | 1 | 125 |
| MUC | 1 | 904 |
| UCLA | 1 | 265 |

COBRE – Center for Biomedical Research Excellence, MCIC – MIND Clinical Imaging Consortium, MIMICSS – Multimodal Imaging in Chronic Schizophrenia Study, MUC – Munich Imaging Database, UCLA – UCLA Consortium for Neuropsychiatric Phenomics LA5c Study

**Table S4.** Detailed neurotransmitter map information incl. tracer, number of subjects, age, sex, and study.

| Neurotransmitter | Tracer | N | Mean Age ± SD | References |
|---|---|---|---|---|
| 5-HT1A | cumi101 | 8 | 28.4 ± 8.8 | (18) |
| 5-HT1B | p943 | 23 | 28.7 ± 7 | (19–26) |
| 5-HT2A | cimbi36 | 29 | 22.6 ± 2.7 | (18) |
| 5-HT4 | sb207145 | 59 | 25.9 ± 5.3 | (18) |
| 5-HT6 | gsk215083 | 30 | 36.6 ± 9.0 | (27,28) |
| 5-HTT | dasb | 100 | 25.1 ± 5.8 | (18) |
| A4B2 | flubatine | 30 | 33.5 ± 10.7 | (20,29,30) |
| CB1 | omar | 77 | 30.0 ± 8.9 | (31–34) |
| COX-1 | ps13 | 10 | 29.3 ± 7.2 | (35,36) |
| D1 | sch23390 | 13 | 33.0 ± 13.0 | (37) |
| D2 | flb457 | 55 | 32.5 ± 9.7 | (20,38–42) |
| DAT | fpcit | 174 | 61.0 ± 11.0 | (43) |
| FDOPA | fluorodopa | 12 | 55.1 ± 16.6 | (44) |
| GABAA | flumazenil | 5 | 43.0 ± 4.0 | (43) |
| GABAA5 | ro154513 | 10 | 25.4 ± 3.2 | (45) |
| H3 | gsk189254 | 8 | 31.7 ± 9.0 | (19,20,46) |
| HDAC | martinostat | 8 | 28.6 ± 7.6 | (47) |
| M1 | lsn3172176 | 24 | 40.5 ± 11.7 | (20,48) |
| mGluR5 | abp688 | 73 | 19.9 ± 3.0 | (20,49) |
| MU | carfentanil | 204 | 32.3 ± 10.8 | (50) |
| NET | mrb | 77 | 33.4 ± 9.2 | (20,51–54) |
| NMDA | ge179 | 29 | 40.9 ± 12.7 | (20,55–57) |
| SV2A | ucbj | 76 | 48.9 ± 18.4 | (32,48,58–70) |
| TSPO | pbr28 | 6 | 57.8 ± 8.1 | (71,72) |
| VAChT | feobv | 18 | 66.8 ± 6.8 | (20,73) |

5-HT1A – Serotonin 1A receptor, 5-HT1B – Serotonin 1B receptor, 5-HT2A – Serotonin 2A receptor, 5-HT4 – Serotonin 4 receptor, 5-HT6 – Serotonin 6 receptor, 5-HTT – Serotonin transporter, A4B2 – Alpha-4 beta-2 nicotinic receptor, CB1 – Cannabinoid receptor 1, COX-1 – Cyclooxygenase 1, D1 – Dopamine D1 receptor, D2 – Dopamine D2 receptor, DAT – Dopamine transporter, FDOPA – Fluorodopa, GABAa – GABA receptor α, GABAA5 – GABA receptor alpha 5, H3 – Histamine H3 receptor, HDAC – Histone deacetylase, M1 – Muscarinic acetylcholine receptor, mGluR5 – Metabotropic glutamate receptor 5, MU – Mu opioid receptor, NET – Norepinephrine transporter, NMDA – NMDA receptor, SV2A – Synaptic vesicle protein 2A, TSPO – Translocator protein, VAChT – Vesicular acetylcholine transporter

**Table S5.** Statistics and corresponding p-values for testing the effectiveness of covariate corrections.

| Model | Site Effect ($X^2$, p) | Age Correlation (r, p) | Sex Difference (W, z, p) |
|---|---|---|---|
| NT | $X^2(7)=5.49$, $p=0.600$ | $r=-0.02$, $p=0.745$ | $W=47383$, $z=-0.11$, $p=0.912$ |
| PCA | $X^2(7)=9.56$, $p=0.215$ | $r=0.0019$, $p=0.969$ | $W=47450$, $z=-0.06$, $p=0.956$ |
| Stacked | $X^2(7)=9.24$, $p=0.236$ | $r=-0.0005$, $p=0.992$ | $W=47020$, $z=-0.41$, $p=0.681$ |

**Table S6.** Estimates and their corresponding standard errors, T-statistics, and FDR-corrected p-values for the estimated marginal means of diagnosis (SCZ, MDD, BD, ADHD, CHR-P, BPD) on decision scores of the NT model, and simple slopes of BrainAGE in each diagnosis (SCZ, MDD, BD, ADHD, CHR-P, BPD) on decision scores of the PCA model.

| | **NT model** | | | | **PCA model** | | | |
|---|---|---|---|---|---|---|---|---|
| | **Estimate** | **SE** | **T** | $p_{FDR}$ | **Estimate** | **SE** | **T** | $p_{FDR}$ |
| SCZ - ADHD | 0.44 | 0.14 | 3.00 | 0.026* | -0.09 | 0.03 | -2.76 | 0.059 |
| SCZ – BD | 0.42 | 0.10 | 4.33 | 0.000* | -0.02 | 0.02 | -0.92 | 1.000 |
| SCZ – BPD | 0.32 | 0.12 | 2.54 | 0.086 | -0.05 | 0.02 | -1.93 | 0.376 |
| SCZ – MDD | 0.27 | 0.11 | 2.56 | 0.086 | 0.00 | 0.02 | 0.17 | 1.000 |
| ADHD – BD | -0.01 | 0.17 | -0.10 | 1.000 | 0.08 | 0.04 | 2.14 | 0.263 |
| ADHD – BPD | -0.12 | 0.18 | -0.65 | 1.000 | 0.04 | 0.04 | 1.10 | 1.000 |
| ADHD – MDD | -0.17 | 0.17 | -1.00 | 1.000 | 0.09 | 0.04 | 2.57 | 0.093 |
| BD – BPD | -0.11 | 0.15 | -0.75 | 1.000 | -0.03 | 0.03 | -1.16 | 1.000 |
| BD – MDD | -0.16 | 0.13 | -1.22 | 1.000 | 0.02 | 0.02 | 0.80 | 1.000 |
| BPD - MDD | -0.05 | 0.15 | -0.33 | 1.000 | 0.05 | 0.03 | 1.73 | 0.501 |

* Significant at $\alpha = 0.05$.

ADHD – Attention Deficit Hyperactivity Disorder, BD – Bipolar Disorder, BPD – Borderline Personality Disorder, MDD – Major Depression, SCZ – Schizophrenia, SE – Standard Error

**Table S7.** Spearman correlations and explained variance ($R^2$) of BrainAGE on model decision scores in each diagnostic group for NT and PCA model.

| | NT model | | | PCA model | | |
|---|---|---|---|---|---|---|
| **Group** | **r** | **$R^2$** | **$p_{FDR}$** | **r** | **$R^2$** | **$p_{FDR}$** |
| ADHD | 0.20 | 0.04 | 0.527 | 0.58 | 0.34 | < 0.001* |
| BD | -0.10 | 0.01 | 0.527 | 0.45 | 0.20 | < 0.001* |
| BPD | 0.25 | 0.06 | 0.287 | 0.47 | 0.22 | < 0.001* |
| MDD | -0.06 | 0.00 | 0.530 | 0.23 | 0.05 | 0.019* |
| SCZ | -0.03 | 0.00 | 0.530 | 0.33 | 0.11 | < 0.001* |

* Significant at $\alpha = 0.05$ after FDR correction.

ADHD – Attention Deficit Hyperactivity Disorder, BD – Bipolar Disorder, BPD – Borderline Personality Disorder, MDD – Major Depression, SCZ – Schizophrenia

**Table S8.** Item-level linear regression model of decision scores of NT and PCA model with PANSS.

| | | | NT model | | | | | PCA model | | | | |
|---|---|---|---|---|---|---|---|---|---|---|---|---|
| **Item** | **Item Description** | **1/VIF** | **Estimate** | **SE** | **T** | ***p*** | **η²** | **Estimate** | **SE** | **T** | ***p*** | **η²** |
| | Intercept | --- | 0.08 | 0.17 | 0.49 | 0.625 | --- | -0.08 | 0.20 | -0.39 | 0.697 | --- |
| P1 | Delusions | 0.28 | -0.09 | 0.05 | -1.70 | 0.090 | 0.00 | -0.06 | 0.06 | -1.05 | 0.296 | 0.01 |
| P2 | Conceptual Disorganization | 0.43 | 0.03 | 0.05 | 0.66 | 0.509 | 0.00 | -0.01 | 0.05 | -0.12 | 0.908 | 0.00 |
| P3 | Hallucinatory Behavior | 0.68 | 0.06 | 0.04 | 1.47 | 0.141 | 0.01 | 0.04 | 0.04 | 0.90 | 0.368 | 0.00 |
| P4 | Excitement | 0.49 | 0.04 | 0.05 | 0.78 | 0.436 | 0.00 | 0.09 | 0.06 | 1.62 | 0.105 | 0.00 |
| P5 | Grandiosity | 0.53 | 0.07 | 0.05 | 1.50 | 0.134 | 0.01 | 0.14 | 0.05 | 2.56 | 0.011* | 0.02 |
| P6 | Suspiciousness/Persecution | 0.37 | 0.02 | 0.05 | 0.39 | 0.697 | 0.00 | 0.02 | 0.06 | 0.30 | 0.765 | 0.00 |
| P7 | Hostility | 0.37 | -0.05 | 0.07 | -0.75 | 0.456 | 0.00 | -0.08 | 0.08 | -1.06 | 0.290 | 0.02 |
| N1 | Blunted Affect | 0.34 | -0.05 | 0.05 | -0.96 | 0.339 | 0.00 | -0.12 | 0.06 | -2.03 | 0.043* | 0.00 |
| N2 | Emotional Withdrawal | 0.25 | 0.06 | 0.06 | 1.00 | 0.320 | 0.01 | 0.10 | 0.07 | 1.54 | 0.125 | 0.02 |
| N3 | Poor Rapport | 0.30 | -0.13 | 0.06 | -2.29 | 0.023* | 0.01 | -0.11 | 0.07 | -1.73 | 0.084 | 0.00 |
| N4 | Passive/Apathetic Withdrawal | 0.30 | -0.02 | 0.05 | -0.46 | 0.644 | 0.00 | 0.01 | 0.06 | 0.13 | 0.894 | 0.01 |
| N5 | Difficulty in Abstract Thinking | 0.56 | 0.03 | 0.04 | 0.68 | 0.494 | 0.00 | 0.09 | 0.05 | 1.90 | 0.058 | 0.02 |
| N6 | Lack of Spontaneity and Flow of Conversation | 0.42 | -0.03 | 0.05 | -0.50 | 0.621 | 0.00 | 0.01 | 0.06 | 0.23 | 0.814 | 0.00 |
| N7 | Stereotyped Thinking | 0.45 | -0.07 | 0.05 | -1.39 | 0.164 | 0.00 | -0.03 | 0.06 | -0.51 | 0.610 | 0.00 |
| G1 | Somatic Concern | 0.58 | 0.03 | 0.04 | 0.74 | 0.460 | 0.01 | -0.03 | 0.04 | -0.58 | 0.562 | 0.00 |
| G2 | Anxiety | 0.39 | 0.01 | 0.05 | 0.11 | 0.911 | 0.00 | 0.08 | 0.06 | 1.36 | 0.176 | 0.00 |
| G3 | Guilt Feelings | 0.48 | -0.06 | 0.04 | -1.39 | 0.165 | 0.00 | -0.04 | 0.05 | -0.85 | 0.396 | 0.01 |
| G4 | Tension | 0.43 | -0.06 | 0.05 | -1.23 | 0.218 | 0.01 | -0.06 | 0.06 | -1.09 | 0.278 | 0.01 |
| G5 | Mannerisms and Posturing | 0.42 | -0.10 | 0.06 | -1.64 | 0.102 | 0.01 | -0.08 | 0.07 | -1.13 | 0.258 | 0.00 |
| G6 | Depression | 0.46 | 0.06 | 0.04 | 1.35 | 0.178 | 0.01 | -0.04 | 0.05 | -0.71 | 0.477 | 0.00 |
| G7 | Motor Retardation | 0.46 | 0.01 | 0.05 | 0.12 | 0.906 | 0.00 | 0.02 | 0.06 | 0.41 | 0.683 | 0.00 |
| G8 | Uncooperativeness | 0.49 | 0.21 | 0.07 | 3.06 | 0.002* | 0.03 | 0.15 | 0.08 | 1.90 | 0.059 | 0.01 |
| G9 | Unusual Thought Content | 0.30 | 0.09 | 0.05 | 1.64 | 0.102 | 0.02 | 0.04 | 0.06 | 0.73 | 0.468 | 0.00 |
| G10 | Disorientation | 0.62 | -0.03 | 0.06 | -0.53 | 0.598 | 0.00 | -0.05 | 0.07 | -0.80 | 0.426 | 0.00 |
| G11 | Poor Attention | 0.48 | -0.03 | 0.05 | -0.49 | 0.628 | 0.00 | 0.01 | 0.06 | 0.09 | 0.930 | 0.00 |
| G12 | Lack of Judgment and Insight | 0.55 | -0.06 | 0.04 | -1.28 | 0.202 | 0.00 | -0.02 | 0.05 | -0.42 | 0.675 | 0.00 |
| G13 | Disturbance of Volition | 0.52 | 0.16 | 0.05 | 3.19 | 0.002* | 0.04 | 0.08 | 0.06 | 1.37 | 0.171 | 0.01 |
| G14 | Poor Impulse Control | 0.45 | -0.09 | 0.05 | -1.84 | 0.066 | 0.01 | -0.15 | 0.06 | -2.55 | 0.011* | 0.02 |
| G15 | Preoccupation | 0.33 | 0.05 | 0.05 | 0.96 | 0.336 | 0.01 | 0.00 | 0.05 | 0.03 | 0.975 | 0.00 |
| G16 | Active Social Avoidance | 0.27 | 0.05 | 0.05 | 1.04 | 0.298 | 0.00 | 0.09 | 0.06 | 1.47 | 0.142 | 0.01 |
| Model Statistics | | | $F(30, 300) = 2.07$, $p = 0.001$, $R^2 = 0.17$ | | | | | $F(30, 300) = 1.77$, $p = 0.010$, $R^2 = 0.15$ | | | | |

* Significant at α = 0.05

PANSS – Positive and Negative Syndrome Scale, VIF – Variance Inflation Factor

**Table S9.** Item-level linear regression model of decision scores of NT and PCA model with SAPS.

| | | | NT model | | | | | PCA model | | | | |
|---|---|---|---|---|---|---|---|---|---|---|---|---|
| **Item** | **Item Description** | **1/VIF** | **Estimate** | **SE** | **T** | ***p*** | **η²** | **Estimate** | **SE** | **T** | ***p*** | **η²** |
| | Intercept | --- | 0.08 | 0.15 | 0.51 | 0.615 | --- | -0.03 | 0.15 | -0.19 | 0.851 | --- |
| 1 | Hallucinations: Auditory | 0.19 | 0.00 | 0.11 | 0.00 | 0.997 | 0.01 | -0.04 | 0.11 | -0.32 | 0.750 | 0.00 |
| 2 | Hallucinations: Voices Commenting | 0.12 | -0.40 | 0.22 | -1.83 | 0.072 | 0.00 | -0.39 | 0.21 | -1.81 | 0.075 | 0.01 |
| 3 | Hallucinations: Voices Conversing | 0.12 | 0.31 | 0.18 | 1.71 | 0.092 | 0.04 | 0.25 | 0.18 | 1.41 | 0.163 | 0.00 |
| 4 | Hallucinations: Somatic or Tactile Hallucinations | 0.28 | -0.14 | 0.17 | -0.80 | 0.425 | 0.03 | 0.12 | 0.17 | 0.69 | 0.492 | 0.00 |
| 5 | Hallucinations: Olfactory Hallucinations | 0.44 | -0.13 | 0.32 | -0.40 | 0.688 | 0.05 | 0.12 | 0.32 | 0.37 | 0.716 | 0.02 |
| 6 | Hallucinations: Visual Hallucinations | 0.39 | -0.10 | 0.14 | -0.72 | 0.472 | 0.00 | -0.08 | 0.13 | -0.63 | 0.528 | 0.00 |
| 8 | Delusions: Persecutory | 0.16 | 0.18 | 0.14 | 1.30 | 0.199 | 0.02 | 0.21 | 0.14 | 1.50 | 0.140 | 0.01 |
| 9 | Delusions: Jealousy | 0.75 | -0.46 | 0.50 | -0.92 | 0.362 | 0.05 | 0.26 | 0.49 | 0.53 | 0.598 | 0.00 |
| 10 | Delusions: Sin or Guilt | 0.39 | -0.02 | 0.21 | -0.07 | 0.941 | 0.00 | 0.04 | 0.21 | 0.18 | 0.856 | 0.00 |
| 11 | Delusions: Grandiose | 0.34 | -0.15 | 0.12 | -1.24 | 0.221 | 0.00 | -0.28 | 0.12 | -2.33 | 0.023* | 0.01 |
| 12 | Delusions: Religious | 0.43 | 0.04 | 0.15 | 0.27 | 0.788 | 0.03 | -0.18 | 0.14 | -1.24 | 0.221 | 0.00 |
| 13 | Delusions: Somatic | 0.14 | -0.37 | 0.29 | -1.29 | 0.203 | 0.00 | -0.57 | 0.28 | -2.02 | 0.048* | 0.00 |
| 14 | Delusions: Ideas of Reference | 0.23 | -0.15 | 0.12 | -1.26 | 0.212 | 0.01 | -0.20 | 0.12 | -1.67 | 0.100 | 0.04 |
| 15 | Delusions: Being Controlled | 0.18 | 0.17 | 0.20 | 0.86 | 0.392 | 0.00 | 0.36 | 0.19 | 1.86 | 0.068 | 0.02 |
| 16 | Delusions: Mind Reading | 0.26 | 0.20 | 0.14 | 1.38 | 0.173 | 0.02 | 0.25 | 0.14 | 1.80 | 0.077 | 0.01 |
| 17 | Thought Broadcasting | 0.12 | -0.04 | 0.33 | -0.13 | 0.898 | 0.00 | -0.39 | 0.32 | -1.22 | 0.227 | 0.02 |
| 18 | Thought Insertion | 0.21 | -0.12 | 0.26 | -0.44 | 0.663 | 0.00 | -0.09 | 0.26 | -0.35 | 0.725 | 0.00 |
| 19 | Thought Withdrawal | 0.15 | 0.26 | 0.43 | 0.60 | 0.552 | 0.02 | 0.34 | 0.42 | 0.80 | 0.425 | 0.03 |
| 21 | Bizarre Behavior: Clothing and Appearance | 0.59 | 0.38 | 0.22 | 1.76 | 0.083 | 0.01 | 0.43 | 0.21 | 2.01 | 0.048* | 0.01 |
| 22 | Bizarre Behavior: Social and Sexual Behavior | 0.21 | -0.21 | 0.19 | -1.12 | 0.266 | 0.00 | -0.34 | 0.18 | -1.86 | 0.067 | 0.02 |
| 23 | Bizarre Behavior: Aggressive and Agitated Behavior | 0.36 | -0.14 | 0.14 | -1.01 | 0.314 | 0.03 | -0.07 | 0.14 | -0.52 | 0.605 | 0.03 |
| 24 | Bizarre Behavior: Repetitive or Stereotyped Behavior | 0.54 | 0.59 | 0.36 | 1.62 | 0.110 | 0.03 | 0.43 | 0.36 | 1.21 | 0.229 | 0.02 |
| 26 | Positive Formal Thought Disorder: Derailment | 0.13 | 0.04 | 0.17 | 0.23 | 0.822 | 0.01 | -0.03 | 0.17 | -0.19 | 0.848 | 0.06 |
| 27 | Positive Formal Thought Disorder: Tangentiality | 0.20 | 0.06 | 0.13 | 0.47 | 0.639 | 0.01 | 0.21 | 0.13 | 1.59 | 0.118 | 0.05 |
| 28 | Positive Formal Thought Disorder: Incoherence | 0.29 | -0.20 | 0.19 | -1.07 | 0.288 | 0.00 | -0.21 | 0.19 | -1.14 | 0.260 | 0.00 |
| 29 | Positive Formal Thought Disorder: Illogicality | 0.27 | 0.15 | 0.21 | 0.71 | 0.480 | 0.00 | 0.23 | 0.20 | 1.16 | 0.252 | 0.01 |

| | | | | | | | | | | | |
|---|---|---|---|---|---|---|---|---|---|---|---|
| 30 | Positive Formal Thought Disorder: Circumstantiality | 0.20 | -0.28 | 0.14 | -1.96 | 0.055 | 0.00 | -0.30 | 0.14 | -2.19 | 0.032* | 0.01 |
| 31 | Positive Formal Thought Disorder: Pressure of speech | 0.34 | 0.23 | 0.13 | 1.77 | 0.081 | 0.05 | 0.18 | 0.13 | 1.44 | 0.155 | 0.03 |
| 32 | Positive Formal Thought Disorder: Distractible speech | 0.61 | 0.21 | 0.16 | 1.37 | 0.177 | 0.03 | 0.20 | 0.15 | 1.29 | 0.202 | 0.02 |
| 33 | Positive Formal Thought Disorder: Clanging | 0.25 | 0.49 | 0.41 | 1.20 | 0.236 | 0.00 | 0.89 | 0.40 | 2.19 | 0.032* | 0.02 |
| 35 | Inappropriate Affect | 0.28 | 0.56 | 0.25 | 2.23 | 0.029* | 0.07 | 0.79 | 0.25 | 3.18 | 0.002* | 0.13 |
| Model Statistics | | | $F(31, 65) = 1.20$, $p = 0.268$, $R^2 = 0.36$ | | | | | $F(31, 65) = 1.32$, $p = 0.176$, $R^2 = 0.39$ | | | | |

* Significant at α = 0.05

SAPS – Scale for the Assessment of Positive Symptoms, VIF – Variance Inflation Factor

**Table S10.** Item-level linear regression model of decision scores of NT and PCA model with SANS.

| | | | NT model | | | | | PCA model | | | | |
|---|---|---|---|---|---|---|---|---|---|---|---|---|
| **Item** | **Item Description** | **1/VIF** | **Estimate** | **SE** | **T** | ***p*** | **η²** | **Estimate** | **SE** | **T** | ***p*** | **η²** |
| | Intercept | --- | 0.02 | 0.16 | 0.12 | 0.903 | --- | 0.12 | 0.19 | 0.61 | 0.541 | --- |
| 1 | Affective Flattening: Unchanging Facial Expression | 0.27 | 0.04 | 0.10 | 0.43 | 0.665 | 0.00 | 0.03 | 0.12 | 0.27 | 0.785 | 0.00 |
| 2 | Affective Flattening: Decreased Spontaneous Movements | 0.31 | 0.02 | 0.11 | 0.17 | 0.862 | 0.00 | 0.04 | 0.13 | 0.27 | 0.786 | 0.00 |
| 3 | Affective Flattening: Paucity of Expressive Gestures | 0.17 | 0.02 | 0.13 | 0.17 | 0.864 | 0.00 | -0.01 | 0.16 | -0.04 | 0.971 | 0.01 |
| 4 | Affective Flattening: Poor Eye Contact | 0.59 | 0.07 | 0.09 | 0.81 | 0.422 | 0.00 | 0.15 | 0.11 | 1.43 | 0.155 | 0.01 |
| 5 | Affective Flattening: Affective Nonresponsivity | 0.29 | -0.08 | 0.10 | -0.78 | 0.436 | 0.01 | -0.04 | 0.13 | -0.36 | 0.722 | 0.00 |
| 6 | Affective Flattening: Lack of Vocal Inflections | 0.26 | 0.03 | 0.10 | 0.29 | 0.769 | 0.00 | -0.22 | 0.13 | -1.79 | 0.075 | 0.02 |
| 8 | Alogia: Poverty of Speech | 0.29 | -0.06 | 0.10 | -0.61 | 0.540 | 0.00 | 0.14 | 0.12 | 1.21 | 0.228 | 0.02 |
| 9 | Alogia: Poverty of Content of Speech | 0.47 | 0.02 | 0.08 | 0.26 | 0.795 | 0.00 | 0.10 | 0.10 | 0.98 | 0.331 | 0.00 |
| 10 | Alogia: Blocking | 0.44 | -0.14 | 0.09 | -1.48 | 0.141 | 0.01 | -0.10 | 0.11 | -0.87 | 0.387 | 0.01 |
| 11 | Alogia: Increased Latency of Response | 0.41 | 0.02 | 0.10 | 0.21 | 0.834 | 0.00 | -0.04 | 0.12 | -0.32 | 0.750 | 0.00 |
| 13 | Avolition-Apathy: Grooming and Hygiene | 0.54 | 0.00 | 0.06 | 0.05 | 0.961 | 0.00 | -0.06 | 0.07 | -0.93 | 0.353 | 0.00 |
| 14 | Avolition-Apathy: Impersistence at Work or School | 0.52 | 0.04 | 0.07 | 0.60 | 0.550 | 0.00 | 0.04 | 0.08 | 0.42 | 0.674 | 0.00 |
| 15 | Avolition-Apathy: Physical Anergia | 0.58 | 0.02 | 0.07 | 0.24 | 0.807 | 0.00 | 0.09 | 0.08 | 1.09 | 0.278 | 0.01 |
| 17 | Anhedonia-Asociality: Recreational Interests and Activities | 0.74 | -0.01 | 0.03 | -0.25 | 0.805 | 0.00 | -0.04 | 0.04 | -1.06 | 0.291 | 0.01 |
| 18 | Anhedonia-Asociality: Sexual Interest and Activity | 0.62 | 0.02 | 0.05 | 0.48 | 0.631 | 0.00 | 0.00 | 0.06 | 0.05 | 0.958 | 0.00 |
| 19 | Anhedonia-Asociality: Ability to Feel Intimacy and Closeness | 0.59 | -0.04 | 0.06 | -0.58 | 0.565 | 0.00 | 0.01 | 0.08 | 0.14 | 0.886 | 0.00 |
| 20 | Anhedonia-Asociality: Relationships with Friends and Peers | 0.50 | 0.03 | 0.06 | 0.42 | 0.673 | 0.00 | 0.05 | 0.07 | 0.72 | 0.471 | 0.00 |
| 22 | Attention: Social Inattentiveness | 0.57 | 0.00 | 0.08 | 0.05 | 0.958 | 0.00 | -0.05 | 0.09 | -0.58 | 0.565 | 0.00 |
| 23 | Attention: Inattentiveness During Mental Status Testing | 0.77 | 0.01 | 0.05 | 0.20 | 0.840 | 0.00 | 0.01 | 0.06 | 0.17 | 0.863 | 0.00 |
| Model Statistics | | | $F(19, 146) = 0.33$, $p = 0.996$, $R^2 = 0.04$ | | | | | $F(19, 146) = 0.84$, $p = 0.661$, $R^2 = 0.10$ | | | | |

SANS – Scale for the Assessment of Negative Symptoms, VIF – Variance Inflation Factor

**Table S11.** Item-level linear regression model of decision scores of NT and PCA model with CDSS.

| | | | NT model | | | | | PCA model | | | | |
|---|---|---|---|---|---|---|---|---|---|---|---|---|
| **Item** | **Item Description** | **1/VIF** | **Estimate** | **SE** | **T** | ***p*** | **η²** | **Estimate** | **SE** | **T** | ***p*** | **η²** |
| | Intercept | --- | 0.15 | 0.17 | 0.91 | 0.365 | --- | 0.12 | 0.20 | 0.62 | 0.539 | -- |
| 1 | Depression | 0.34 | 0.58 | 0.29 | 2.02 | 0.048* | 0.06 | 0.26 | 0.33 | 0.79 | 0.430 | 0.01 |
| 2 | Hopelessness | 0.32 | 0.07 | 0.28 | 0.24 | 0.813 | 0.00 | -0.17 | 0.33 | -0.53 | 0.601 | 0.00 |
| 3 | Self-Depreciation | 0.55 | -0.09 | 0.19 | -0.46 | 0.648 | 0.01 | 0.15 | 0.22 | 0.67 | 0.508 | 0.01 |
| 4 | Guilty Ideas Of Reference | 0.38 | 0.19 | 0.24 | 0.79 | 0.431 | 0.00 | -0.05 | 0.27 | -0.18 | 0.855 | 0.00 |
| 5 | Pathological Guilt | 0.42 | -0.37 | 0.27 | -1.38 | 0.174 | 0.03 | -0.05 | 0.31 | -0.16 | 0.877 | 0.00 |
| 6 | Morning Depression | 0.55 | 0.54 | 0.26 | 2.06 | 0.044* | 0.05 | 0.42 | 0.30 | 1.38 | 0.172 | 0.02 |
| 7 | Early Wakening | 0.62 | 0.33 | 0.24 | 1.35 | 0.182 | 0.01 | 0.25 | 0.28 | 0.89 | 0.379 | 0.01 |
| 8 | Suicide | 0.35 | -0.13 | 0.34 | -0.38 | 0.708 | 0.01 | 0.19 | 0.39 | 0.48 | 0.632 | 0.00 |
| 9 | Observed Depression | 0.30 | -0.79 | 0.35 | -2.27 | 0.027* | 0.08 | -0.59 | 0.41 | -1.46 | 0.151 | 0.03 |
| Model Statistics | | | $F(9, 61) = 1.73$, $p = 0.101$, $R^2 = 0.20$ | | | | | $F(9, 61) = 0.60$, $p = 0.796$, $R^2 = 0.08$ | | | | |

* Significant at α = 0.05

CDSS – Calgary Depression Scale for Schizophrenia, VIF – Variance Inflation Factor

**Table S12.** Item-level linear regression model of decision scores of NT and PCA model with HDRS-17.

| | | | NT model | | | | | PCA model | | | | |
|---|---|---|---|---|---|---|---|---|---|---|---|---|
| **Item** | **Item Description** | **1/VIF** | **Estimate** | **SE** | **T** | ***p*** | **η²** | **Estimate** | **SE** | **T** | ***p*** | **η²** |
| | Intercept | --- | 0.14 | 0.13 | 1.07 | 0.287 | --- | -0.01 | 0.15 | -0.09 | 0.931 | --- |
| 1 | Depressed Mood | 0.29 | 0.03 | 0.08 | 0.32 | 0.752 | 0.00 | 0.07 | 0.09 | 0.70 | 0.482 | 0.00 |
| 2 | Feelings of Guilt | 0.69 | -0.02 | 0.08 | -0.21 | 0.834 | 0.00 | 0.09 | 0.09 | 0.99 | 0.326 | 0.00 |
| 3 | Suicide | 0.57 | -0.03 | 0.07 | -0.45 | 0.651 | 0.00 | 0.02 | 0.08 | 0.21 | 0.834 | 0.00 |
| 4 | Insomnia Early | 0.66 | -0.12 | 0.09 | -1.37 | 0.172 | 0.01 | 0.00 | 0.10 | 0.04 | 0.965 | 0.00 |
| 5 | Insomnia Middle | 0.54 | 0.04 | 0.09 | 0.40 | 0.686 | 0.00 | -0.05 | 0.10 | -0.49 | 0.621 | 0.00 |
| 6 | Insomnia Late | 0.64 | -0.05 | 0.09 | -0.51 | 0.614 | 0.00 | -0.11 | 0.10 | -1.05 | 0.293 | 0.00 |
| 7 | Work and Activities | 0.41 | 0.02 | 0.07 | 0.29 | 0.774 | 0.00 | -0.06 | 0.08 | -0.76 | 0.447 | 0.00 |
| 8 | Psychomotor Retardation | 0.60 | 0.06 | 0.12 | 0.50 | 0.618 | 0.00 | 0.11 | 0.13 | 0.80 | 0.426 | 0.00 |
| 9 | Psychomotor Agitation | 0.82 | -0.05 | 0.08 | -0.72 | 0.473 | 0.01 | -0.08 | 0.09 | -0.95 | 0.345 | 0.01 |
| 10 | Anxiety Psychic | 0.48 | 0.03 | 0.07 | 0.39 | 0.693 | 0.00 | -0.05 | 0.08 | -0.70 | 0.486 | 0.00 |
| 11 | Anxiety Somatic | 0.56 | -0.06 | 0.08 | -0.84 | 0.402 | 0.01 | 0.01 | 0.09 | 0.11 | 0.911 | 0.00 |
| 12 | Somatic Symptoms Gastrointestinal | 0.49 | 0.07 | 0.15 | 0.45 | 0.653 | 0.00 | 0.22 | 0.17 | 1.32 | 0.189 | 0.00 |
| 13 | Somatic Symptoms General | 0.59 | -0.04 | 0.09 | -0.44 | 0.662 | 0.00 | 0.06 | 0.11 | 0.57 | 0.569 | 0.00 |
| 14 | Genital Symptoms | 0.63 | 0.09 | 0.10 | 0.89 | 0.377 | 0.00 | -0.04 | 0.11 | -0.38 | 0.702 | 0.00 |
| 15 | Hypochondriasis | 0.67 | -0.08 | 0.08 | -0.99 | 0.323 | 0.01 | 0.02 | 0.09 | 0.21 | 0.833 | 0.00 |
| 16 | Loss of Weight | 0.62 | -0.04 | 0.10 | -0.40 | 0.692 | 0.00 | -0.11 | 0.11 | -1.04 | 0.302 | 0.01 |
| 17 | Insight | 0.71 | -0.07 | 0.19 | -0.34 | 0.731 | 0.00 | -0.13 | 0.22 | -0.59 | 0.557 | 0.00 |
| Model Statistics | | | $F(17, 197) = 0.54$, $p = 0.932$, $R^2 = 0.04$ | | | | | $F(17, 197) = 0.47$, $p = 0.963$, $R^2 = 0.04$ | | | | |

HDRS-17 – Hamilton Depression Rating Scale, VIF – Variance Inflation Factor

**Table S13.** Detailed information regarding numbers of drug-naïve and exposed patients, coefficients, their corresponding semi-partial R², standard errors, T-statistics, and p-values for the linear regression models.

| | | | NT model | | | | | PCA model | | | | |
|---|---|---|---|---|---|---|---|---|---|---|---|---|
| | **Medication** | **N**$_{exposed}$ (no/yes) | **Estimate (η²)** | | ***SE*** | ***t*** | ***p*** | **Estimate (η²)** | | ***SE*** | ***t*** | ***p*** |
| **COBRE** | Intercept | --- | -0.01 | (- - -) | 0.44 | -0.02 | 0.981 | 0.32 | (- - -) | 0.46 | 0.70 | 0.487 |
| | AP | 6/65 | 0.36 | (0.01) | 0.47 | 0.77 | 0.443 | -0.25 | (0.00) | 0.49 | -0.51 | 0.610 |
| | AT | 54/17 | -0.16 | (0.01) | 0.31 | -0.52 | 0.603 | 0.20 | (0.00) | 0.33 | 0.60 | 0.553 |
| | BZP | 68/3 | 0.02 | (0.00) | 0.65 | 0.03 | 0.979 | 0.21 | (0.00) | 0.68 | 0.31 | 0.757 |
| | MS | 66/5 | 0.36 | (0.01) | 0.51 | 0.71 | 0.479 | 1.28 | (0.08) | 0.53 | 2.41 | 0.019* |
| **MCIC** | Intercept | --- | 0.52 | (- - -) | 0.37 | 1.39 | 0.166 | -0.45 | (- - -) | 0.49 | -0.94 | 0.351 |
| | AP | 5/97 | -0.18 | (0.00) | 0.38 | -0.47 | 0.642 | 1.09 | (0.05) | 0.50 | 2.20 | 0.030 |
| **MIMICSS** | Intercept | --- | 0.70 | (- - -) | 0.55 | 1.26 | 0.213 | 1.05 | (- - -) | 0.89 | 1.18 | 0.243 |
| | AP | 3/66 | -0.61 | (0.02) | 0.56 | -1.09 | 0.281 | -0.70 | (0.01) | 0.89 | -0.79 | 0.434 |
| | AT | 57/11 | 0.08 | (0.00) | 0.27 | 0.32 | 0.750 | -0.04 | (0.00) | 0.43 | -0.08 | 0.934 |
| | BZP | 55/13 | -0.03 | (0.00) | 0.24 | -0.11 | 0.911 | 0.39 | (0.02) | 0.39 | 1.02 | 0.313 |
| | MS | 67/1 | 0.21 | (0.00) | 0.81 | 0.27 | 0.792 | 0.36 | (0.00) | 1.29 | 0.28 | 0.780 |
| **MUC** | Intercept | --- | 0.06 | (- - -) | 0.40 | 0.14 | 0.890 | -0.18 | (- - -) | 0.44 | -0.42 | 0.683 |
| | AP (CPZ) | 0/24 | 0.00 | (0.15) | 0.00 | 1.69 | 0.108 | 0.00 | (0.12) | 0.00 | 1.58 | 0.131 |
| | AT | 21/3 | -0.66 | (0.03) | 0.68 | -0.97 | 0.346 | -0.01 | (0.00) | 0.76 | -0.02 | 0.985 |
| | BZP | 23/1 | -1.00 | (0.04) | 1.12 | -0.89 | 0.382 | -0.73 | (0.02) | 1.24 | -0.59 | 0.564 |
| | MS | 22/2 | -0.68 | (0.03) | 0.82 | -0.83 | 0.419 | -0.60 | (0.02) | 0.91 | -0.66 | 0.515 |
| **UCLA** | Intercept | --- | -0.23 | (- - -) | 0.10 | -2.18 | 0.031* | -0.27 | (- - -) | 0.13 | -2.17 | 0.032* |
| | AP | 70/69 | 0.42 | (0.09) | 0.15 | 2.83 | 0.005* | 0.35 | (0.04) | 0.18 | 1.90 | 0.059 |
| | AT | 95/44 | 0.19 | (0.01) | 0.16 | 1.21 | 0.229 | 0.19 | (0.01) | 0.19 | 1.00 | 0.317 |
| | BZP | 116/23 | 0.16 | (0.00) | 0.20 | 0.82 | 0.413 | 0.03 | (0.00) | 0.24 | 0.11 | 0.912 |
| | MS | 96/43 | -0.04 | (0.00) | 0.16 | -0.28 | 0.778 | -0.01 | (0.00) | 0.19 | -0.08 | 0.940 |

* Significant at α = 0.05.

AP – Antipsychotics, AT – Antidepressants, BZP – Benzodiazepines, CPZ – Chlorpromazine equivalent dose, MS – Mood stabilizers

**Table S14.** Internal validation and stability measures for kmeans, pam, hierarchical (average and Ward linkage), and agnes (average and Ward linkage) with 2 to 10 clusters.

| Clustering algorithm | Measure | | 2 | 3 | 4 | 5 | 6 | 7 | 8 | 9 | 10 |
|---|---|---|---|---|---|---|---|---|---|---|---|
| k-means | Internal Validation | APN | 0.02 | 0.20 | 0.10 | 0.27 | 0.09 | 0.10 | 0.14 | 0.17 | 0.18 |
| | | AD | 69.7 | 68.8 | 64.5 | 64.5 | 60.5 | 59.4 | 59.2 | 59.0 | 58.2 |
| | | ADM | 1.47 | 11.25 | 5.36 | 15.56 | 5.00 | 5.43 | 7.21 | 9.51 | 9.69 |
| | | FOM | 6.86 | 6.50 | 6.28 | 6.26 | 6.15 | 6.08 | 6.07 | 5.99 | 5.92 |
| | Stability | Connectivity | 110.6 | 234.7 | 365.9 | 302.9 | 307.3 | 337.1 | 341.8 | 364.0 | 348.4 |
| | | Dunn | 0.13 | 0.11 | 0.07 | 0.08 | 0.11 | 0.11 | 0.11 | 0.11 | 0.11 |
| | | Silhouette | 0.23 | 0.18 | 0.13 | 0.14 | 0.13 | 0.14 | 0.13 | 0.13 | 0.14 |
| PAM | Internal Validation | APN | 0.05 | 0.06 | 0.10 | 0.08 | 0.09 | 0.08 | 0.08 | 0.07 | 0.12 |
| | | AD | 70.5 | 67.6 | 65.7 | 63.5 | 62.3 | 60.8 | 59.5 | 58.1 | 58.0 |
| | | ADM | 3.34 | 2.84 | 4.60 | 3.84 | 4.26 | 3.75 | 3.62 | 3.15 | 5.65 |
| | | FOM | 6.77 | 6.59 | 6.41 | 6.33 | 6.24 | 6.15 | 6.10 | 6.03 | 5.99 |
| | Stability | Connectivity | 210.0 | 353.9 | 425.0 | 419.5 | 460.6 | 486.2 | 458.0 | 473.2 | 499.3 |
| | | Dunn | 0.08 | 0.08 | 0.09 | 0.11 | 0.11 | 0.10 | 0.10 | 0.10 | 0.11 |
| | | Silhouette | 0.20 | 0.12 | 0.10 | 0.11 | 0.10 | 0.10 | 0.10 | 0.11 | 0.09 |
| hierarchical (average linkage) | Internal Validation | APN | 0.00 | 0.00 | 0.00 | 0.00 | 0.00 | 0.00 | 0.00 | 0.01 | 0.03 |
| | | AD | 78.1 | 77.9 | 77.6 | 76.2 | 76.1 | 75.8 | 75.6 | 75.4 | 75.2 |
| | | ADM | 0.07 | 0.14 | 0.36 | 0.26 | 0.60 | 0.57 | 0.69 | 0.74 | 1.84 |
| | | FOM | 7.21 | 7.20 | 7.13 | 7.07 | 7.06 | 7.04 | 7.03 | 7.01 | 6.96 |
| | Stability | Connectivity | 2.9 | 5.9 | 11.5 | 33.5 | 39.4 | 45.2 | 45.2 | 45.2 | 51.2 |
| | | Dunn | 0.38 | 0.38 | 0.31 | 0.25 | 0.25 | 0.25 | 0.25 | 0.25 | 0.20 |
| | | Silhouette | 0.48 | 0.38 | 0.34 | 0.30 | 0.27 | 0.25 | 0.25 | 0.22 | 0.12 |
| hierarchical (Ward linkage) | Internal Validation | APN | 0.12 | 0.16 | 0.22 | 0.23 | 0.22 | 0.19 | 0.21 | 0.22 | 0.22 |
| | | AD | 72.4 | 68.8 | 67.0 | 65.4 | 63.6 | 61.7 | 60.9 | 60.2 | 59.1 |
| | | ADM | 8.89 | 9.03 | 12.41 | 14.34 | 13.08 | 10.45 | 11.42 | 12.20 | 11.44 |
| | | FOM | 6.80 | 6.61 | 6.53 | 6.45 | 6.33 | 6.25 | 6.20 | 6.15 | 6.07 |
| | Stability | Connectivity | 77.3 | 115.0 | 143.7 | 169.5 | 198.5 | 218.0 | 227.8 | 241.1 | 245.7 |
| | | Dunn | 0.14 | 0.14 | 0.14 | 0.13 | 0.16 | 0.15 | 0.15 | 0.15 | 0.15 |
| | | Silhouette | 0.15 | 0.12 | 0.14 | 0.09 | 0.11 | 0.12 | 0.11 | 0.11 | 0.11 |
| agnes | | APN | 0.00 | 0.00 | 0.00 | 0.00 | 0.00 | 0.00 | 0.00 | 0.01 | 0.03 |

| | | | | | | | | | | | |
|---|---|---|---|---|---|---|---|---|---|---|---|
| (average linkage) | Internal Validation | AD | 78.1 | 77.9 | 77.6 | 76.2 | 76.1 | 75.8 | 75.6 | 75.4 | 75.2 |
| | | ADM | 0.07 | 0.14 | 0.36 | 0.26 | 0.60 | 0.57 | 0.69 | 0.74 | 1.84 |
| | | FOM | 7.21 | 7.20 | 7.13 | 7.07 | 7.06 | 7.04 | 7.03 | 7.01 | 6.96 |
| | Stability | Connectivity | 2.9 | 5.9 | 11.5 | 33.5 | 39.4 | 45.2 | 45.2 | 45.2 | 51.2 |
| | | Dunn | 0.38 | 0.38 | 0.31 | 0.25 | 0.25 | 0.25 | 0.25 | 0.25 | 0.20 |
| | | Silhouette | 0.48 | 0.38 | 0.34 | 0.30 | 0.27 | 0.25 | 0.25 | 0.22 | 0.12 |
| agnes (Ward linkage) | Internal Validation | APN | 0.19 | 0.19 | 0.21 | 0.19 | 0.17 | 0.14 | 0.17 | 0.17 | 0.15 |
| | | AD | 73.4 | 69.1 | 67.0 | 64.9 | 62.9 | 60.9 | 60.5 | 59.6 | 58.2 |
| | | ADM | 15.43 | 11.98 | 12.97 | 11.21 | 10.70 | 7.70 | 9.81 | 10.74 | 8.38 |
| | | FOM | 6.85 | 6.66 | 6.49 | 6.39 | 6.29 | 6.21 | 6.17 | 6.11 | 6.01 |
| | Stability | Connectivity | 51.4 | 98.7 | 134.2 | 156.4 | 190.2 | 211.6 | 225.6 | 234.0 | 238.6 |
| | | Dunn | 0.14 | 0.14 | 0.14 | 0.18 | 0.17 | 0.14 | 0.14 | 0.14 | 0.14 |
| | | Silhouette | 0.15 | 0.12 | 0.13 | 0.14 | 0.11 | 0.11 | 0.12 | 0.11 | 0.11 |

agnes – Agglomerative Nesting, PAM – Partitioning Around Medoids, APN – Average Proportion of Non-overlap, AD – Average Distance, ADM – Average Distance between Means, FOM – Figure of Merit, Dunn – Dunn Index, Silhouette – Silhouette Width

**Table S15.** Distribution of clusters across diagnostic groups.

| Diagnostic group | N Cluster 1 (%) | | N Cluster 2 (%) | |
|---|---|---|---|---|
| ADHD | 9 | (3.80%) | 10 | (2.01%) |
| BD | 12 | (5.06%) | 32 | (6.44%) |
| BPD | 11 | (4.64%) | 24 | (4.83%) |
| HC | 118 | (49.8%) | 192 | (38.6%) |
| MDD | 14 | (5.91%) | 37 | (7.44%) |
| SCZ | 73 | (30.8%) | 202 | (40.6%) |
| Total | 237 | | 497 | |

ADHD – Attention Deficit Hyperactivity Disorder, BD – Bipolar Disorder, BPD – Borderline Personality Disorder, MDD – Major Depression, SCZ – Schizophrenia

**Table S16.** Comparison of cortical and subcortical correlations between clusters including mean differences, statistics, FDR-corrected p-value, and effect size r.

| | Cortical correlations | | | | | Subcortical correlations | | | | |
|---|---|---|---|---|---|---|---|---|---|---|
| **NT** | **Mean Difference** | ***W*** | ***z*** | $p_{FDR}$ | $r_z$ | **Mean Difference** | ***W*** | ***z*** | $p_{FDR}$ | $r_z$ |
| 5-HT1A | 1.90 | 99724 | 4.70 | 0.000* | 0.17 | 16.07 | 126072 | 14.51 | 0.000* | 0.54° |
| 5-HT1B | -1.40 | 82258 | -1.80 | 0.087 | -0.07 | 1.76 | 104093 | 6.33 | 0.000* | 0.23 |
| 5-HT2A | 0.87 | 91974 | 1.82 | 0.087 | 0.07 | 0.44 | 93522 | 2.39 | 0.023* | 0.09 |
| 5-HT4 | 2.45 | 113541 | 9.84 | 0.000* | 0.36° | 19.67 | 131619 | 16.58 | 0.000* | 0.61° |
| 5-HT6 | 0.54 | 91774 | 1.74 | 0.095 | 0.06 | 5.01 | 114097 | 10.05 | 0.000* | 0.37° |
| 5-HTT | -0.53 | 82426 | -1.74 | 0.095 | -0.06 | 2.05 | 102129 | 5.60 | 0.000* | 0.21 |
| A4B2 | 3.06 | 116777 | 11.05 | 0.000* | 0.41° | -1.60 | 79868 | -2.69 | 0.010* | -0.10 |
| CB1 | -2.05 | 82049 | -1.88 | 0.077 | -0.07 | 4.20 | 99933 | 4.78 | 0.000* | 0.18 |
| COX-1 | 3.34 | 99187 | 4.50 | 0.000* | 0.17 | -3.88 | 62104 | -9.30 | 0.000* | -0.34° |
| D1 | -1.45 | 73583 | -5.03 | 0.000* | -0.19 | -1.81 | 73843 | -4.93 | 0.000* | -0.18 |
| D2 | 3.12 | 115954 | 10.74 | 0.000* | 0.40° | -27.87 | 39463 | -17.73 | 0.000* | -0.65° |
| DAT | 0.39 | 91138 | 1.50 | 0.147 | 0.06 | -11.85 | 48514 | -14.36 | 0.000* | -0.53° |
| FDOPA | 4.19 | 125616 | 14.34 | 0.000* | 0.53° | 5.19 | 105796 | 6.96 | 0.000* | 0.26 |
| GABAA | -0.25 | 85502 | -0.59 | 0.576 | -0.02 | 17.41 | 123884 | 13.70 | 0.000* | 0.51° |
| GABAA5 | -0.50 | 86382 | -0.27 | 0.790 | -0.01 | 5.81 | 119095 | 11.91 | 0.000* | 0.44° |
| H3 | 3.11 | 103367 | 6.06 | 0.000* | 0.22 | -0.87 | 85038 | -0.77 | 0.472 | -0.03 |
| HDAC | 0.01 | 87844 | 0.28 | 0.790 | 0.01 | 16.45 | 117854 | 11.45 | 0.000* | 0.42° |
| M1 | -0.96 | 82782 | -1.61 | 0.123 | -0.06 | 5.57 | 122091 | 13.03 | 0.000* | 0.48° |
| mGluR5 | 2.69 | 102659 | 5.79 | 0.000* | 0.21 | -26.73 | 43892 | -16.09 | 0.000* | -0.59° |
| MU | -0.60 | 81990 | -1.90 | 0.075 | -0.07 | -5.63 | 71683 | -5.74 | 0.000* | -0.21 |
| NET | 0.53 | 90005 | 1.08 | 0.303 | 0.04 | -0.60 | 81407 | -2.12 | 0.046* | -0.08 |
| NMDA | 3.51 | 109884 | 8.48 | 0.000* | 0.31° | 1.58 | 94993 | 2.94 | 0.005* | 0.11 |
| SV2A | 2.22 | 100886 | 5.13 | 0.000* | 0.19 | 9.71 | 121370 | 12.76 | 0.000* | 0.47° |
| TSPO | -1.07 | 78341 | -3.26 | 0.002* | -0.12 | 3.63 | 112321 | 9.39 | 0.000* | 0.35° |
| VAChT | 1.44 | 94220 | 2.65 | 0.011* | 0.10 | 6.66 | 122024 | 13.00 | 0.000* | 0.48° |

* Significant at α = 0.05 after FDR correction.

° Effect size $r_z$ |≥| 0.3 (medium effect size).

5-HT1A – Serotonin 1A receptor, 5-HT1B – Serotonin 1B receptor, 5-HT2A – Serotonin 2A receptor, 5-HT4 – Serotonin 4 receptor, 5-HT6 – Serotonin 6 receptor, 5-HTT – Serotonin transporter, A4B2 – Alpha-4 beta-2 nicotinic receptor, CB1 – Cannabinoid receptor 1, COX-1 – Cyclooxygenase 1, D1 – Dopamine D1 receptor, D2 – Dopamine D2 receptor, DAT – Dopamine transporter, FDOPA – Fluorodopa, GABAa – GABA receptor α, GABAA5 – GABA receptor alpha 5, H3 –

Histamine H3 receptor, HDAC – Histone deacetylase, M1 – Muscarinic acetylcholine receptor, mGluR5 – Metabotropic glutamate receptor 5, MU – Mu opioid receptor, NET – Norepinephrine transporter, NMDA – NMDA receptor, SV2A – Synaptic vesicle protein 2A, TSPO – Translocator protein, VAChT – Vesicular acetylcholine transporter

**Table S17.** Linear model coefficients (Estimates, SEs, *t*-values, *p*-values, and η²) for associations between symptom dimensions (SAPS, SANS, PANSS) and distance to centroid within each cluster.

| | | Cluster 1 | | | | | Cluster 2 | | | | |
|---|---|---|---|---|---|---|---|---|---|---|---|
| | | **Estimate** | **SE** | **T** | ***p*** | **η²** | **Estimate** | **SE** | **T** | ***p*** | **η²** |
| SAPS (global) | Intercept | 65.24 | 6.37 | 10.24 | 0.000* | --- | 52.68 | 2.43 | 21.69 | 0.000* | --- |
| | Hallucinations | -2.54 | 2.33 | -1.09 | 0.285 | 0.01 | 0.56 | 1.02 | 0.55 | 0.586 | 0.01 |
| | Delusions | 3.15 | 2.08 | 1.51 | 0.142 | 0.08 | -0.36 | 0.98 | -0.37 | 0.713 | 0.00 |
| | Bizzare Behavior | -1.61 | 3.21 | -0.50 | 0.620 | 0.01 | 1.47 | 1.27 | 1.16 | 0.249 | 0.02 |
| | Positive Formal Thought Disorder | -1.77 | 2.61 | -0.68 | 0.504 | 0.01 | 0.27 | 1.19 | 0.23 | 0.818 | 0.00 |
| | Model statistics | *F*(4, 26) = 0.87, *p* = 0.497, R² = 0.12 | | | | | *F*(4, 85) = 0.64, *p* = 0.633, R² = 0.03 | | | | |
| SANS (global) | Intercept | 54.50 | 9.33 | 5.84 | 0.000 | --- | 49.12 | 2.83 | 17.33 | 0.000 | --- |
| | Affective Flattening | 4.52 | 2.80 | 1.62 | 0.115 | 0.02 | -1.51 | 1.07 | -1.41 | 0.161 | 0.03 |
| | Alogia | -6.42 | 3.07 | -2.09 | 0.044* | 0.12 | -2.59 | 1.18 | -2.19 | 0.031* | 0.02 |
| | Avolition Apathy | 3.17 | 3.25 | 0.98 | 0.336 | 0.03 | 1.03 | 1.15 | 0.89 | 0.373 | 0.03 |
| | Anhedonia Asociality | -1.80 | 3.55 | -0.51 | 0.616 | 0.01 | 0.92 | 1.16 | 0.80 | 0.428 | 0.01 |
| | Attention | 2.26 | 2.19 | 1.03 | 0.309 | 0.03 | 1.05 | 1.01 | 1.04 | 0.302 | 0.01 |
| | Model statistics | *F*(5, 34) = 1.65, *p* = 0.173, R² = 0.20 | | | | | *F*(5, 110) = 2.37, *p* = 0.044, R² = 0.10 | | | | |
| PANSS (Wallwork) | Intercept | 48.69 | 6.15 | 7.91 | 0.000* | --- | 42.03 | 3.09 | 13.59 | 0.000* | --- |
| | Positive | -1.73 | 1.60 | -1.08 | 0.285 | 0.03 | 0.77 | 1.02 | 0.76 | 0.451 | 0.01 |
| | Negative | -0.86 | 1.50 | -0.58 | 0.566 | 0.02 | -0.85 | 0.91 | -0.93 | 0.352 | 0.00 |
| | Disorganized/ Concrete | 0.65 | 1.81 | 0.36 | 0.721 | 0.00 | 0.37 | 1.22 | 0.30 | 0.762 | 0.00 |
| | Excited | 2.53 | 2.69 | 0.94 | 0.353 | 0.02 | -0.56 | 1.30 | -0.44 | 0.664 | 0.00 |
| | Depressed | -0.24 | 1.71 | -0.14 | 0.888 | 0.00 | 1.16 | 0.92 | 1.27 | 0.208 | 0.01 |
| | Model statistics | *F*(5, 51) = 0.48, *p* = 0.790, R² = 0.04 | | | | | *F*(5, 141) = 0.65, *p* = 0.662, R² = 0.02 | | | | |

* Significant at α = 0.05.

PANSS – Positive and Negative Syndrome Scale, SANS – Scale for the Assessment of Negative Symptoms, SAPS – Scale for the Assessment of Positive Symptoms

**Table S18.** Spearman correlation coefficients for illness duration and HDRS-17 total score with distance to centroid in cluster 1 and cluster 2.

| | **Cluster 1** | | | | **Cluster 2** | | | |
|---|---|---|---|---|---|---|---|---|
| | **N** | **r** | **$R^2$** | ***p*** | **N** | **r** | **$R^2$** | ***p*** |
| Illness Duration | 36 | 0.27 | 0.07 | 0.102 | 105 | -0.10 | 0.01 | 0.300 |
| HDRS-17 (total) | 30 | -0.27 | 0.07 | 0.143 | 76 | -0.27 | 0.07 | 0.017* |

* Significant at α = 0.05.

HDRS-17 – Hamilton Depression Rating Scale

**Table S19.** Detailed information on differences between clusters regarding illness duration, PANSS factors, SAPS and SANS global ratings, and total scores of CDSS, and HDRS-17.

| Scale | Item | $N_{Cluster1}$/ $N_{Cluster2}$ | Mean Difference | W | z | $p_{FDR}$ | $r_z$ |
|---|---|---|---|---|---|---|---|
| Illness Duration | | 37/107 | -1.88 | 2530 | -0.70 | 0.487 | -0.06 |
| BrainAGE | | 237/497 | 1.25 | 92953 | 2.18 | 0.029* | 0.08 |
| PANSS factors (Wallwork) | Positive | 58/148 | 0.10 | 6231 | 0.59 | 0.922 | 0.04 |
| | Negative | 58/148 | 0.13 | 6298 | 0.77 | 1.000 | 0.05 |
| | Disorganized/concrete | 58/147 | 0.08 | 6196 | 0.58 | 0.701 | 0.04 |
| | Excited | 57/148 | -0.16 | 5757 | -0.30 | 0.701 | -0.02 |
| | Depressed | 58/148 | 0.22 | 6461 | 1.19 | 1.000 | 0.08 |
| SAPS global | Hallucinations | 31/90 | -0.28 | 1746 | -0.88 | 0.600 | -0.08 |
| | Delusions | 31/90 | 0.42 | 2089 | 1.20 | 0.600 | 0.11 |
| | Bizzare Behavior | 31/90 | 0.02 | 1967 | 0.53 | 0.600 | 0.05 |
| | Positive Formal Thought Disorder | 31/90 | 0.18 | 2005 | 0.73 | 0.600 | 0.07 |
| SANS global | Affective Flattening | 40/117 | 0.25 | 3345 | 0.77 | 0.575 | 0.06 |
| | Alogia | 40/118 | 0.29 | 3430 | 1.06 | 0.575 | 0.08 |
| | Avolition Apathy | 40/118 | 0.38 | 3490 | 1.26 | 0.930 | 0.10 |
| | Anhedonia Asociality | 40/117 | 0.02 | 3139 | -0.08 | 0.575 | -0.01 |
| | Attention | 40/118 | 0.28 | 3362 | 0.75 | 0.575 | 0.06 |
| CDSS | Total | 11/32 | 0.45 | 222 | -0.54 | 0.591 | -0.08 |
| HDRS-17 | Total | 31/78 | 0.18 | 1733 | 0.19 | 0.851 | 0.02 |

* Significant at α = 0.05.

CDSS – Calgary Depression Scale for Schizophrenia, HDRS-17 – HDRS-17 – Hamilton Depression Rating Scale
